# Association of Healthy Dietary Patterns and Cardiorespiratory Fitness in the Community

**DOI:** 10.1101/2023.02.09.23285714

**Authors:** Michael Y. Mi, Priya Gajjar, Maura E. Walker, Patricia Miller, Vanessa Xanthakis, Venkatesh L. Murthy, Martin G. Larson, Ramachandran S. Vasan, Ravi V. Shah, Gregory D. Lewis, Matthew Nayor

## Abstract

**Aims:** To evaluate the associations of dietary indices and quantitative CRF measures in a large, community-based sample harnessing metabolomic profiling to interrogate shared biology.

**Methods:** Framingham Heart Study (FHS) participants underwent maximum effort cardiopulmonary exercise tests for CRF quantification (via peak VO_2_) and completed semi-quantitative FFQs. Dietary quality was assessed by the Alternative Healthy Eating Index (AHEI) and Mediterranean-style Diet Score (MDS), and fasting blood concentrations of 201 metabolites were quantified.

**Results:** In 2380 FHS participants (54±9 years, 54% female, BMI 28±5 kg/m^2^), 1-SD higher AHEI and MDS were associated with 5.1% (1.2 ml/kg/min, p<0.0001) and 4.4% (1.0 ml/kg/min, p<0.0001) greater peak VO_2_ in linear models adjusted for age, sex, total energy intake, cardiovascular risk factors, and physical activity. In participants with metabolite profiling (N=1154), 24 metabolites were concordantly associated with both dietary indices and peak VO_2_ in multivariable-adjusted linear models (FDR<5%). These metabolites included C6 and C7 carnitines, C16:0 ceramide, and dimethylguanidino valeric acid, which were higher with lower CRF and poorer dietary quality and are known markers of insulin resistance and cardiovascular risk. Conversely, C38:7 phosphatidylcholine plasmalogen and C38:7 and C40:7 phosphatidylethanolamine plasmalogens were associated with higher CRF and favorable dietary quality and may link to lower cardiometabolic risk.

**Conclusion:** Higher diet quality is associated with greater CRF cross-sectionally in a middle-aged community-dwelling sample, and metabolites highlight potential shared favorable effects on health.

## Introduction

Cardiorespiratory fitness (CRF) is an integrative measure of physiological reserve capacity and metabolic health that is closely linked with cardiovascular outcomes [1]. While an individual’s CRF may be partially determined by standard cardiovascular disease (CVD) risk factors, there remains a large amount of interindividual variability in CRF that is not well understood [2]. Lifestyle factors are also likely to contribute to CRF and may present easily modifiable avenues for improving CRF [3]. Healthy dietary patterns are an important component of cardiovascular health [4], but it remains uncertain whether they are also related to CRF. Indeed, while putative mechanisms linking a healthy diet and improved fitness have been proposed to include anti-inflammatory effects or increased availability of antioxidants, these mechanisms are incompletely elucidated [5].

Although association of diet with CRF has been previously reported, prior studies relied primarily on treadmill exercise time as a surrogate for peak CRF, focused on specific (often younger) populations, and used varying dietary assessments that focused on specific food types rather than standardized and integrated indices of evidence-based healthy patterns of eating [5–14]. Furthermore, few studies have examined the association of macronutrient intake with fitness in population-based cohorts. One observational study of well-educated White adults presenting for preventive cardiac evaluation suggested that lower fat and higher carbohydrate intake as percentages of total energy were associated with higher fitness [13]. Whether such a relation holds in community-dwelling individuals consuming more contemporary diets requires further investigation.

To address these knowledge gaps, we related indices of healthy dietary patterns in community-dwelling individuals from the Framingham Heart Study (FHS) with maximum effort cardiopulmonary exercise test (CPET) measures including peak VO_2_ (the “gold standard” assessment of CRF) and complementary CRF measures. We then further explored the relations between healthy dietary patterns and peak VO_2_ by comparing their joint associations with >200 circulating blood metabolites. Our overarching objective was to quantify the cross-sectional relation of healthy dietary patterns with CRF measures and to gain insight into potential mechanisms through metabolite profiling. We hypothesized that healthier patterns of eating would associate with greater CRF and that metabolites associated with dietary quality and CRF would highlight potential pathways linking diet and CRF.

## Methods

### Study sample

Descriptions of recruitment and enrollment of the FHS Generation Three, Omni Generation Two, and New Offspring cohorts are reported [15,16]. Briefly, these cohorts were recruited together and attended their first and second study visits in 2002 to 2005 and 2008 to 2011, respectively. Of the 3521 study participants attending the third study visit (2016 to 2019), 3117 participants underwent maximum effort CPET and 2494 of these individuals also completed semi-quantitative FFQs. We excluded individuals with missing CPET measures (N=18), inadequate volitional effort (peak RER <1.05; N=105), and missing covariate information (N=9), resulting in a final analytical sample of 2380 individuals (**Supplementary Figure 1**). From this subsample, targeted metabolomics profiling by liquid chromatography-mass spectrometry (LC-MS) was performed on peripheral blood drawn after ≥8 hours of fasting in 1154 individuals (**Supplementary Figure 1**). Protocols were approved by the Institutional Review Boards at Boston University Medical Campus and Massachusetts General Hospital and all participants provided written informed consent.

### Covariate assessment

We included the following covariates in our analysis: sex, age, total energy intake, BMI, smoking status, total cholesterol, HDL, systolic blood pressure (SBP), self-reported use of anti-hypertensive medication, diabetes, and physical activity index (PAI). Total energy intake was calculated as described below from FFQ. We classified smoking status as never, former, or current (within the last year). Diabetes was assessed as fasting blood glucose ≥126 mg/dL, non-fasting blood glucose ≥200 mg/dL, or use of glucose lowering medications. PAI was calculated based on a weighted estimate of total oxygen consumption from questionnaires of typical daily activities. For sensitivity analysis, we substituted PAI with objectively measured sedentary time and amount of moderate-vigorous physical activity (MVPA) as measured by accelerometers worn for up to eight days, as described previously [3].

### Dietary patterns

Dietary intake was assessed using the Harvard semi-quantitative FFQ, which quantifies intake during the last year of 126 dietary items ranging from never or less than once per month to ≥6 servings/day. The FFQ has demonstrated good reproducibility and validity compared to a one-week dietary record [17]. We excluded FFQs with >12 blank items or with improbable estimated daily energy intake, defined as <600 or ≥4000 kcal/day for females and <600 or ≥4200 kcal/day for males. For each participant, macronutrient intake was estimated by summing frequency of consumption multiplied by nutrient composition for the specific serving size of all food items on the FFQ [18]. Total energy intake was estimated based on 4 kcal/g for carbohydrates and proteins and 9 kcal/g for fats, and percent macronutrient intake was calculated by dividing each component’s energy contribution by the total energy.

We calculated indices of adherence to two dietary patterns, the Alternative Health Eating Index (AHEI) and the Mediterranean-style Diet Score (MDS), both of which are associated with CVD outcomes [19,20]. The AHEI assigns a score of 0 to 10 for each of the 11 dietary components and, thus, has a total range of 0 to 110 with higher scores indicating greater adherence [19]. The AHEI components include a higher intake of vegetables, fruits, whole grains, nuts/legumes, omega-3 fatty acids, PUFA, and a lower intake of sugar-sweetened beverages/fruit juice, red/processed meat, trans fatty acids, sodium, and sex-specific moderate alcohol use (**Supplementary Table 1**). To calculate the MDS score, we assigned cohort- and sex-specific quartile scores (score of 0 for the lowest quartile and 3 for the highest quartile) to each of eight components: vegetables, fruits, whole grains, nuts, legumes, red meat (scored in reverse), fish, and the ratio of MUFA to SFA, and a single score for sex-specific moderate alcohol use [21]. The MDS ranged from 0 to 25 with higher scores indicating greater similarity to a Mediterranean-style diet (**Supplementary Table 1**).

**Table 1:**
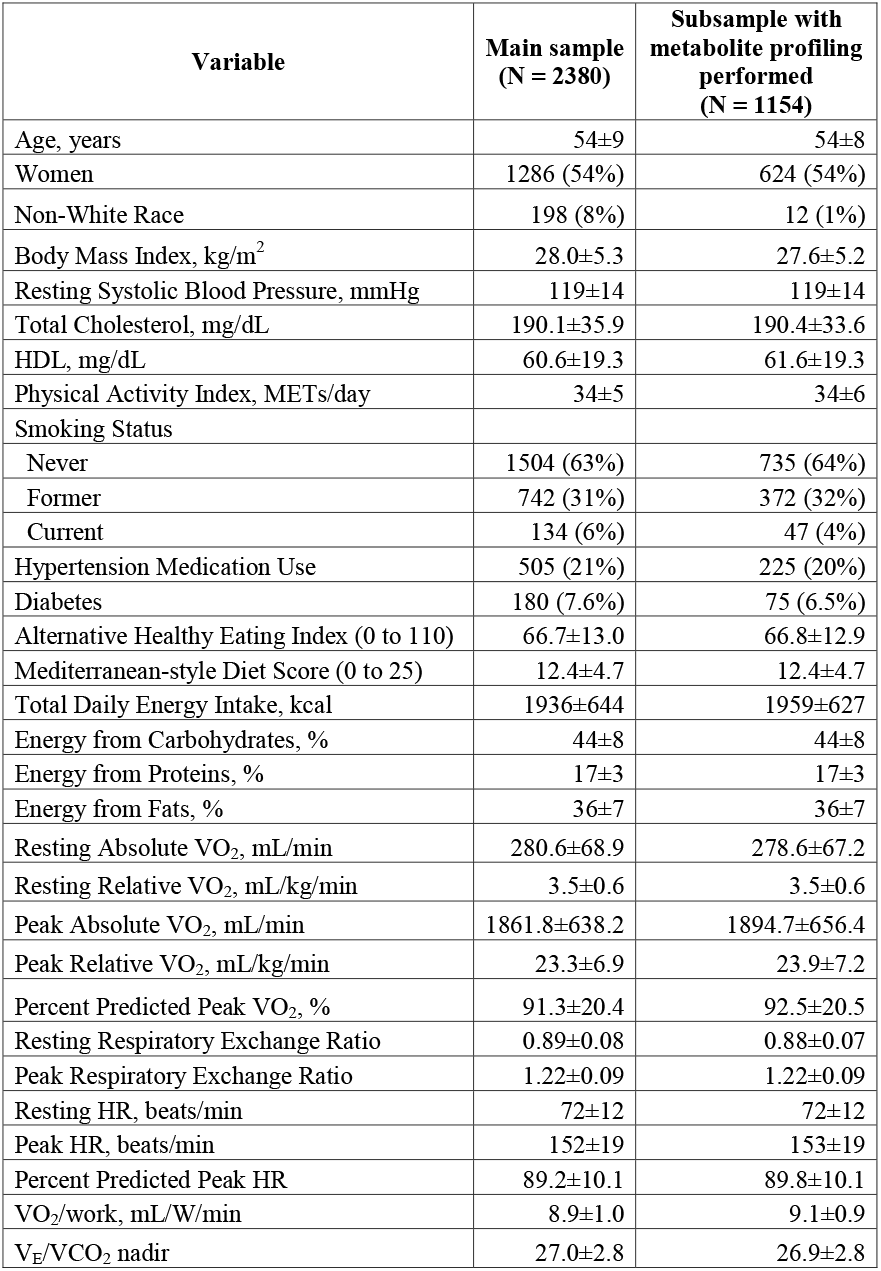
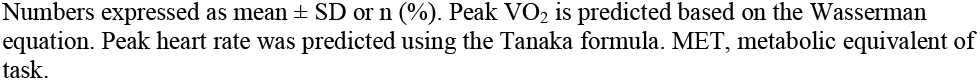
Sample characteristics.

### Cardiopulmonary exercise testing

Details of the CPET protocol and data collection have been described [3,22]. All participants exercised on the same cycle ergometer (Lode, Groningen, Netherlands) with breath-by-breath gas exchange values measured by the same metabolic cart (MedGraphics, St. Paul, MN). The assessment included at least 3 minutes of resting gas exchange measures, followed by 3 minutes of unloaded exercise, and then incremental ramp exercise on one of two (15 or 25 W/min) ramp protocols [3]. Peak VO_2_ was assessed as the highest 30-second median during the final minute of exercise. The following gas exchange and exercise variables were included: resting absolute VO_2_, resting relative VO_2_ (absolute VO_2_ divided by body weight), peak absolute VO_2_, peak relative VO_2_, percent predicted peak absolute VO_2_ by the Wasserman equation, resting RER, peak RER, resting HR, peak HR, percent predicted peak HR by the Tanaka formula, VO_2_/work relationship, and ventilatory efficiency (minute ventilation per 1 L/min of carbon dioxide expiration, assessed as V_E_/VCO_2_ nadir) [23].

### Metabolite profiling

Fasting plasma samples were collected, immediately centrifuged, and stored at -80 L without freeze-thaw cycles before assaying. Targeted measurement of polar metabolites was performed on a Nexera X2 UHPLC (Shimadzu Corporation, Kyoto, Japan) liquid chromatograph coupled to a Q Exactive hybrid quadrupole Orbitrap mass spectrometer (Thermo Fisher Scientific, Waltham, MA) with samples injected on a 150×2 mm, 3-μm Atlantis hydrophilic interaction liquid chromatography (HILIC) column (Waters Corporation, Milford, MA) [22].

Peak integration was performed using TraceFinder (Thermo Fisher Scientific) and identified a broad selection of metabolites based on reference samples. These metabolite classes included amino acids, amino acid metabolites, acylcarnitines, purines, pyrimidines, glycerophospholipids, and sphingolipids.

### Statistical analysis

Peak VO_2_ (absolute and relative) were natural log transformed for improved normality and CPET measures and dietary indices were standardized to mean=0, SD=1 to facilitate comparison and interpretation. Metabolite levels were inverse rank-normalized for analysis and missing (below detection limit) metabolite values were imputed to half of the lowest detected values [22].

We evaluated the cross-sectional associations of dietary indices (AHEI and MDS; independent variables) with CPET fitness measures (dependent variables) using multivariable linear regression. First, we adjusted for age, sex, and total energy intake (model 1). In model 2, we additionally adjusted for BMI, smoking status, total cholesterol, HDL, SBP, hypertension medication use, diabetes, and PAI. We tested for effect modification by age (<54 years [median age] vs. ≥54 years), sex, and BMI (<25, 25–30, or ≥30 kg/m^2^) on the associations between dietary patterns and key CPET variables (peak relative VO_2_, resting relative VO_2_, and percent predicted peak VO_2_) using multiplicative interaction terms. In secondary analyses, we used the same multivariable linear regression models above to relate deciles of the percent daily energy intake from protein, fat, and carbohydrate with CPET fitness measures. A Benjamini-Hochberg false discovery rate (FDR) of 5% was used to determine statistical significance within each CPET variable and diet index analysis.

We conducted sensitivity analyses for the main associations of AHEI and MDS with CPET variables by substituting objective measures of sedentary time and MVPA for PAI in the analytic subset with available accelerometry data. We also examined the interrelationship between resting HR and peak VO_2_ by adjusting each model for the other measure. We evaluated for non-linearity and threshold effects for the relations of the dietary indices with log-transformed relative peak VO_2_ by constructing generalized additive models that applied a smooth function to allow the AHEI or MDS scores to have non-linear relations with log-transformed peak VO_2_; models were adjusted for age, sex, total energy intake, BMI, smoking status, total cholesterol, HDL, SBP, hypertension medication use, diabetes, and PAI. Model fits of the linear and general additive models were compared using ANOVA.

Transformed metabolite levels were then separately related to each of AHEI, MDS, and relative peak VO_2_ using multivariable linear regression. Models were first adjusted for age, sex, total energy intake, and BMI, and then we additionally adjusted for smoking status, total cholesterol, HDL, SBP, hypertension medication use, diabetes, and PAI. An FDR of 5% was used to determine statistical significance separately for associations of metabolites with AHEI, MDS, and peak VO_2_. We then identified metabolites significantly associated with AHEI or MDS total scores and relative peak VO_2_. We related these significant metabolites to individual components of AHEI and MDS using multivariable regression, adjusted for age, sex, total energy intake, BMI, smoking status, cholesterol, HDL, SBP, hypertension medication use, diabetes, and PAI. Again, an FDR of 5% was used to determine statistical significance separately for associations of metabolites with each dietary pattern component. Analyses were performed in R 4.0.3 (R Foundation for Statistical Computing, Vienna, Austria).

## Results

### Sample characteristics

Characteristics of the main study sample and the subsample with available metabolite measures are shown in **Table 1**. Participants had a mean age of 54±9 years, BMI of 28.0±5.3 kg/m^2^, and peak VO_2_ of 23.3±6.9 ml/kg/min (91±20% predicted), and 54% were female. The mean total energy intake was 1849 kcal/day for females and 2039 kcal/day for males. Across the cohort, overall energy intake comprised of approximately 44% carbohydrates, 36% fats, and 17% proteins. Mean AHEI and MDS scores were 66.7±13.0 and 12.4±4.7 respectively, which are largely consistent with ranges reported in other community-based studies [19,24–26].

Spearman correlation between AHEI and MDS was 0.72. Characteristics of the analytic subsample with metabolite profiling performed were similar to the main sample with a higher proportion of White participants.

### Relations of dietary quality assessment with fitness measures

We related two different dietary quality indices (AHEI and MDS) with exercise response measures reflecting distinct physiologies. In models adjusted for age, sex, and total energy intake, higher dietary quality (both higher AHEI and MDS) was associated with higher resting VO_2_ and with global CRF as measured by peak VO_2_ (absolute, relative, and percent predicted). Dietary quality was also related to multi-dimensional exercise measures reflecting favorable autonomic function (higher peak HR and lower resting HR), greater oxygen uptake kinetics during exercise (VO_2_/work), and favorable central cardiac and pulmonary vascular function (lower V_E_/VCO_2_; **Supplementary Table 2**). With further adjustment for clinical risk factors and potential confounders, we observed attenuation of the relations with resting VO_2_, but associations with exercise measures remained robust (**Table 2**) with broad consistency across the two dietary indices. In the multivariable adjusted models, a 1-SD higher AHEI and MDS were associated with 5.1% (4.2% to 5.9%) and 4.4% (3.5% to 5.3%) greater peak relative VO_2_, respectively (which translates to 1.2 and 1.0 ml/kg/min at the sample mean of 23.3 ml/kg/min). We verified the linearity of these associations with general additive models, and no threshold effects were observed (ANOVA p>0.05 comparing linear to general additive models; **Figure 1**).

**Figure 1:**
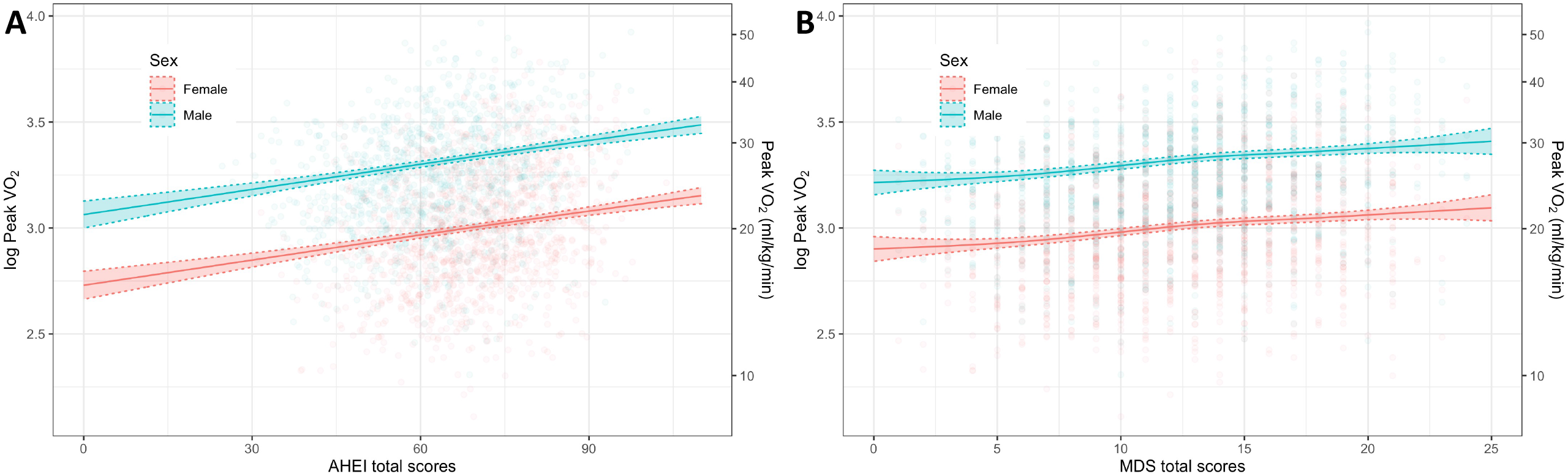
Scatterplots of peak oxygen uptake (VO_2_) vs. Alternative Healthy Eating Index (AHEI) in Panel A and Mediterranean-style Diet Score (MDS) in Panel B stratified by sex and overlaid with fitted curves from general additive models and their 95% confidence bounds. Models were adjusted for age, sex, total energy intake, BMI, smoking status, total cholesterol, HDL, systolic blood pressure, hypertension medication use, diabetes, and physical activity index.

**Table 2:** Cross sectional associations of dietary patterns and cardiopulmonary exercise testing variables.

| CPET Variables | Alternative Healthy Eating Index<br>(N = 2380) |  |  | Mediterranean-style Diet Score<br>(N = 2380) |  |  |
| --- | --- | --- | --- | --- | --- | --- |
|  | Coefficient | SE | FDR p | Coefficient | SE | FDR p |
| Resting Absolute VO<sub>2</sub> | 0.005 | 0.014 | 0.78 | 0.016 | 0.014 | 0.31 |
| Resting Relative VO<sub>2</sub> | 0.005 | 0.019 | 0.80 | 0.020 | 0.019 | 0.31 |
| Peak Absolute VO<sub>2</sub> | 0.147 | 0.013 | <0.0001 | 0.127 | 0.014 | <0.0001 |
| Peak Relative VO<sub>2</sub> | 0.170 | 0.014 | <0.0001 | 0.148 | 0.015 | <0.0001 |
| Percent Predicted Peak VO<sub>2</sub> | 0.212 | 0.020 | <0.0001 | 0.183 | 0.021 | <0.0001 |
| Resting RER | -0.029 | 0.022 | 0.23 | -0.017 | 0.023 | 0.44 |
| Peak RER | -0.032 | 0.021 | 0.17 | -0.036 | 0.022 | 0.13 |
| Resting HR | -0.131 | 0.021 | <0.0001 | -0.115 | 0.022 | <0.0001 |
| Peak HR | 0.055 | 0.018 | 0.0046 | 0.069 | 0.019 | 0.0004 |
| Percent Predicted Peak HR | 0.060 | 0.020 | 0.0049 | 0.077 | 0.021 | 0.0004 |
| VO<sub>2</sub>/work | 0.142 | 0.020 | <0.0001 | 0.113 | 0.020 | <0.0001 |
| V<sub>E</sub>/VCO<sub>2</sub> nadir | -0.089 | 0.020 | <0.0001 | -0.064 | 0.020 | 0.0026 |

Absolute and relative VO<sub>2</sub> at rest and peak were log transformed. Coefficients represent change in CPET variables per one SD increase in dietary scores. Models were adjusted for age, sex, total energy intake, BMI, smoking status, cholesterol, HDL, systolic blood pressure, hypertension medication use, and physical activity index. P-values were adjusted for false discovery rate using the Benjamini-Hochberg method separately for each dietary pattern. CPET, cardiopulmonary exercise testing; FDR, false discovery rate; HR, heart rate; RER, respiratory exchange ratio.

In sensitivity analyses, we substituted objectively measured physical activity and sedentary time (available in a subset of N=1777 individuals) for the questionnaire based PAI as adjustment variables, with minimal changes observed (**Supplementary Table 3**). To further explore whether the relation of a lower resting HR may be partially explained by higher fitness levels, we included both measures in models (**Supplementary Table 4**). Adjustment for peak relative VO_2_ attenuated the effect size of dietary quality’s relations with resting HR, but they remained significant in the main model. Adjustment for resting HR had minimal influence on the effect size of the relations between dietary quality and peak VO_2_.

**Table 3:** Summary of metabolites associated with dietary patterns and peak relative VO_2_.

| Metabolite Class | Metabolites | AHEI/<br>MDS | VO<sub>2</sub> | Diet and Biological Relationships |
| --- | --- | --- | --- | --- |
| Glycerolipids | C34:1 DAG or TAG fragment | ↓ | ↓ | Intermediates of fatty acid metabolism, DAGs are found in small quantities (< 10%) in various seed oils [59], and ingestion has been associated with decreased weight, waist circumference, and serum triglyceride [59] |
| Glycerophosphocholines | C20:1 LPC | ↑ | ↑ | LPCs produced by cleavage of PCs by phospholipase A2, and saturated and shorter LPCs are pro-inflammatory whereas unsaturated and longer LPCs are anti-inflammatory [60,61]. C22:6 LPC associated with lower risk of type 2 diabetes. C22:6 LPC positively associated with AHEI and MDS [24], fish intake [32], and vegetable intake [62], and it increases with fish oil supplementation [33]. C24:0 LPC is inversely associated with diet high in meat and fast food [62]. |
|  | C22:6 LPC | ↑ | ↑ |  |
|  | C24:0 LPC | ↑ | ↑ |  |
| Glycerophosphocholines | C38:7 PC<br>Plasmalogen | ↑ | ↑ | Phospholipids found in neuronal and cardiac cell membranes. Exhibit antioxidant properties and inversely associated with obesity, diabetes, metabolic syndrome [31]; C38:7 PC and C38:7 PE plasmalogens are positively associated with MDS and fish intake [32,39]. Ether C38:7 PE and ether C40:7 PE increases with fish oil supplementation [33]. |
| Glycerophosphoethanolamines | C38:7 PE<br>Plasmalogen<br>C40:7 PE<br>Plasmalogen | ↑ | ↑ |  |
| Ceramides | C16:0 Ceramide<br>d18:1 | ↓ | ↓ | Associated with incident CVD [29], and inversely associated with DASH diet [43]; Elevated level leads to insulin resistance, and lower levels protect from obesity in mouse models [52]. |
| Sphingomyelins | C18:1 SM | ↓ | ↓ | Higher levels associated with diabetes [51]; inversely associated with AHEI, MDS, DASH diet, and fish intake [24,25,43] |
| Fatty esters | C5:1 Carnitine | ↑ | ↑ | Short chain acylcarnitine associated positively with insulin sensitivity [63] |
|  | C6 Carnitine<br>C7 Carnitine | ↓ | ↓ | Medium chain acylcarnitines associated with obesity, type 2 diabetes [28], and CVD [50]; inversely associated with MDS [39] |
| Benzenes | Trimethylbenzene | ↑ | ↑ | Used as industrial solvents and has three isomers, some of which are found naturally in plants [64] |
| Pyrimidines | Ectoine | ↑ | ↑ | Produced by various bacteria species, binds water molecules, and an osmoprotectant [65]; positively associated with chicken intake [32] |
| Amino acids, peptides, and derivatives | Asparagine | ↑ | ↑ | Non-essential amino acid, associated with decreased cardiovascular and all-cause mortality [66,67] |
|  | Cinnamoyl-glycine | ↑ | ↑ | Conjugate of cinnamic acid and glycine and associated with decreased risk of diabetes [68]; cinnamic acid is synthesized in plants, commonly found in cinnamon, and derivatives of cinnamic acid linked with improved glucose homeostasis and insulin resistance [57]. |
|  | N-monomethyl-arginine (Targinine) | ↑ | ↑ | Methylated form of arginine, inhibitor of nitric oxide synthase, and potent vasoconstrictor [54]; associated with decreased risk of incident coronary heart disease in African Americans [56]. |
|  | N-acetyl-ornithine | ↑ | ↑ | Intermediate metabolite in the biosynthesis of arginine; positively associated with tea intake, DASH diet, vegetable intake, and AHEI [32,40,43] |
|  | N-acetyl-tryptophan | ↑ | ↑ | Inhibitor of cytochrome c release and the binding of substance P to neurokinin 1 receptor with potential downstream neuroprotective effects [69] |
|  | Pantothenate (Vitamin B<sub>5</sub>) | ↑ | ↑ | Component of coenzyme A synthesis, essential for fatty acid metabolism; positively associated with MDS [24] |
|  | Pipecolic acid | ↑ | ↑ | Intermediate metabolite in the metabolism of lysine to 2-aminoadipic acid, which is associated with increased risk of incident diabetes [70]; elevated in peroxisome diseases, reported to have an inhibitory effect on the central nervous system [71]; positively associated with MDS and DASH diet [39,43] |
|  | Valine | ↑ | ↑ | A branched chain amino acid, related to increased risk of incident diabetes and insulin resistance [72,73]; metabolized to beta-amino-isobutyric acid, which induces brown adipocyte specific gene expression [74] |
| Carboximidic acids | N-carbamoyl-beta-alanine (Ureidopropionic acid) | ↑ | ↑ | Intermediate metabolite of uracil catabolism to beta-alanine |
| Keto acids | Dimethyl-guanidino valeric acid (DMGV) | ↓ | ↓ | Associated with incident diabetes, nonalcoholic fatty liver disease, coronary artery disease, and cardiovascular mortality [30,58]; positively associated with sugar-sweetened beverage intake and inversely associated with vegetable intake [30] |

AHEI, Alternative Healthy Eating Index; DAG, diacylglycerol; DASH, Dietary Approaches to Stop Hypertension; LPC, lysophosphatidylcholine; MDS, Mediterranean-style Diet Score; PC, phosphatidylcholine; PE, phosphatidylethanolamine; TAG, triacylglycerol. Number after C denotes carbon atoms in acyl side chains and number after colon denotes number of double bonds. References 59 to 74 are included in the Supplementary Material.

Next, we tested for effect modification by age, sex, or BMI on the relation of dietary quality and peak VO_2_ using multiplicative interaction terms. We observed a statistically significant interaction of age with both scores for the association with percent predicted peak VO_2_, such that the effect estimate was higher in individuals younger than the median age of 54 years (**Supplementary Table 5**). There was no evidence of effect modification by sex or BMI.

Finally, we evaluated whether macronutrient components may be related to fitness measures. Consistent with known physiology and prior studies [27], we observed higher carbohydrate intake to be associated with a higher resting RER (**Supplementary Table 6**). Additionally, we observed that after controlling for total daily energy intake, a one decile higher percent energy intake from carbohydrates was associated with lower peak VO_2_, whereas a higher percent energy intake from fat was associated with higher peak VO_2_.

### Shared associations of diet and fitness measures with the circulating metabolome

To evaluate the joint relations of dietary patterns and higher CRF with blood metabolites, we evaluated linear models adjusted for clinical risk factors. In multivariable adjusted models, 83 metabolites were associated with AHEI, 81 with MDS, and 53 with peak relative VO_2_ (FDR <5% for all; **Supplementary Table 7**). Of significant metabolite associations, 67 were shared between the two dietary patterns, 32 were shared between AHEI and peak relative VO_2_, 24 were shared between MDS and peak relative VO_2_, and 24 were common to all three (**Supplementary Figure 2**). Directions of associations of metabolites with favorable dietary quality and with higher peak VO_2_ were consistent **(Figure 2**).

**Figure 2:**
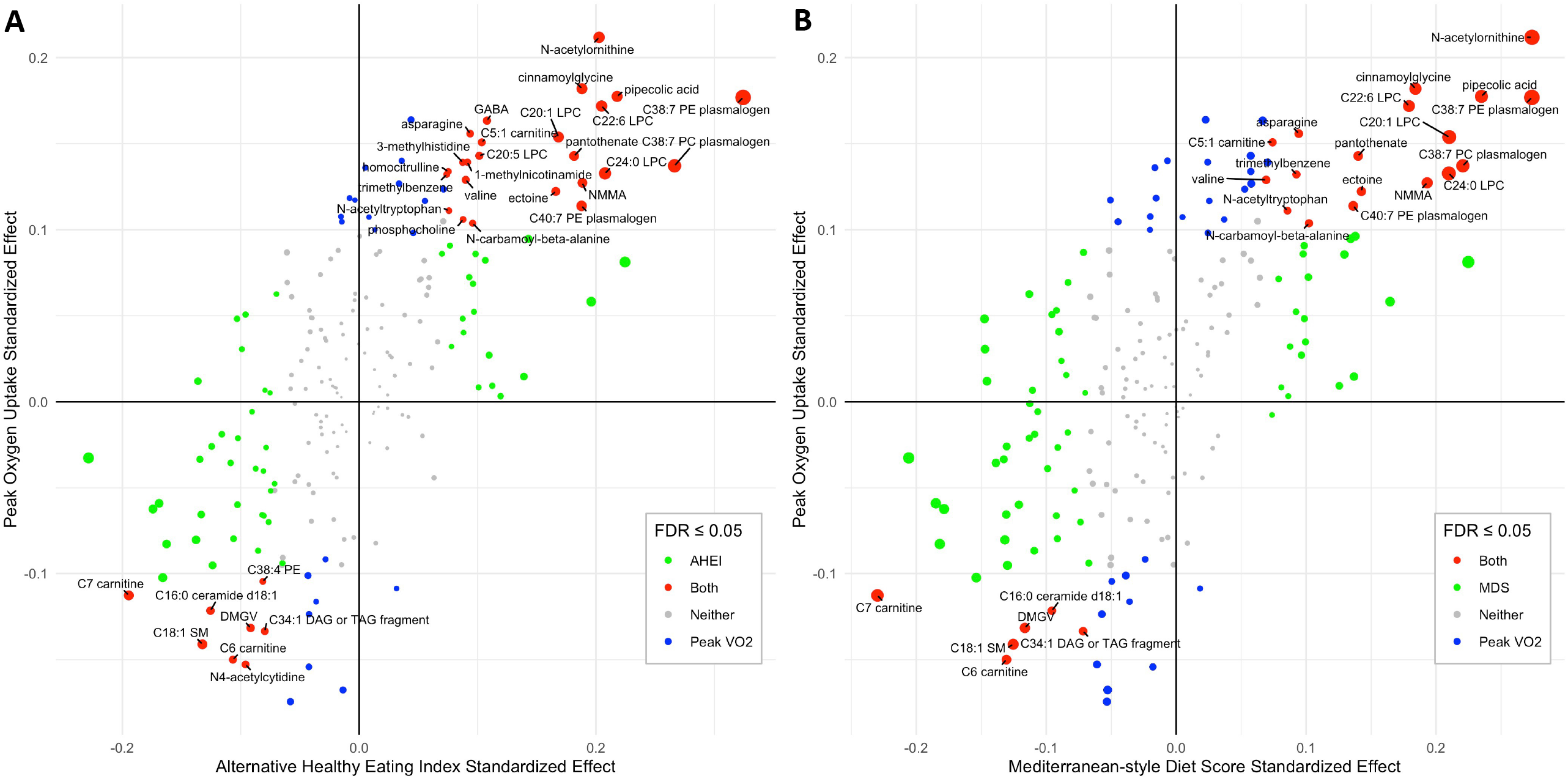
The beta coefficients were plotted for each metabolite in relation to peak oxygen uptake (VO_2_) (y-axis) and dietary indices scores (x-axis) in linear models adjusted for age, sex, total energy intake, BMI, smoking status, total cholesterol, HDL, systolic blood pressure, hypertension medication use, diabetes, and physical activity index. Panel A shows results for Alternative Healthy Eating Index, and Panel B shows results for Mediterranean-style Diet Score. Point sizes reflect the average –log10 p-values for associations between VO_2_ and the respective dietary index. Labeled metabolites have significant associations with respective dietary index and VO_2_ after false discovery rate adjustment. DMGV, dimethylguanidino valeric acid; GABA, gamma-aminobutyric acid; NMMA, LPC, lysophosphatidylcholine; LPE, lysophosphatidylethanolamine; N-monomethyl-arginine; PC, phosphatidylcholine; PE, phosphatidylethanolamine; SM, sphingomyelin. For acyl group nomenclature, the number after C denotes the number of carbon atoms, and the number after the colon denotes the number of double bonds.

**Table 3** summarizes the 24 metabolites associated with both dietary patterns and peak relative VO_2_. These include metabolites previously implicated in cardiometabolic health and fitness as well as novel metabolites with less well-elucidated roles in fitness. Many metabolites have established associations with cardiometabolic disease such as C6 and C7 carnitines, C16:0 ceramide, and dimethylguanidino valeric acid (DMGV), whose higher levels are linked to higher risks of diabetes and/or CVD [28–30] and associated with lower CRF and poorer dietary quality in our cohort. Conversely, higher C38:7 phosphatidylcholine (PC) plasmalogen and C38:7 and C40:7 phosphatidylethanolamine (PE) plasmalogen levels were associated with higher CRF and better dietary quality; as a group, plasmalogens are hypothesized to have antioxidant properties and negatively associated with metabolic diseases [31].

We then further explored the relations of metabolites with individual components of each dietary index (**Figure 3**). Some of the strongest metabolite associations with individual food groups included glycerophospholipids such as C40:7 and C38:7 PE plasmalogens, C38:7 PC plasmalogen, C22:6 lysophosphatidylcholine (LPC), C20:5 LPC, and C38:4 PE, which were positively associated with fish and omega-3 fatty acid intake, consistent with prior work [32,33]. These findings further support the validity of our dietary assessments by demonstrating relations with biochemical signatures of their intake.

**Figure 3:**
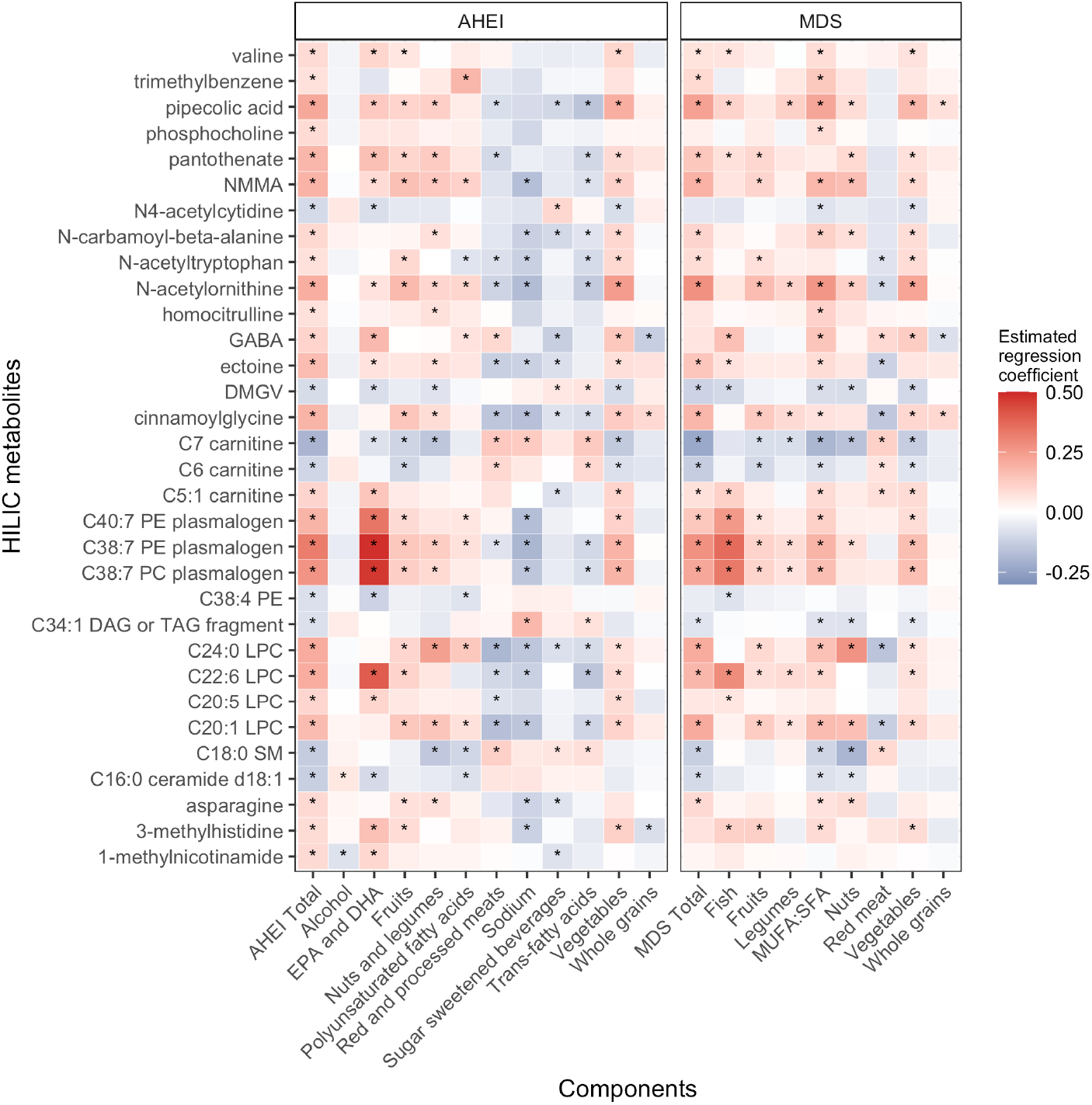
Heat maps of the estimated regression coefficients from models relating metabolites that were significantly related to peak oxygen uptake (VO_2_) and Alternative Healthy Eating Index (AHEI) and Mediterranean-style Diet Score (MDS) to dietary index components are displayed. Linear regression models adjusted for age, sex, total energy intake, BMI, smoking status, total cholesterol, HDL, hypertension medication use, diabetes, and physical activity index. HILIC, hydrophilic interaction liquid chromatography; NMMA, N-monomethyl-arginine; GABA, gamma-aminobutyric acid; DMGV, dimethylguanidino valeric acid; PE, phosphatidylethanolamine; PC, phosphatidylcholine; LPC, lysophosphatidylcholine; LPE, lysophosphatidylethanolamine; SM, sphingomyelin; EPA, eicosapentaenoic acid; DHA, docosahexaenoic acid; MUFA, monounsaturated fatty acid; SFA, saturated fatty acid. For acyl group nomenclature, the number after C denotes the number of carbon atoms, and the number after the colon denotes the number of double bonds.

## Discussion

Here, we provide a comprehensive examination of the associations of healthy dietary patterns with fitness measures in a large, community-based sample and quantify a broad circulating metabolome to explore joint associations of diet and fitness. Two readily applicable healthy dietary patterns were associated with CRF in our study with broad consistency across age, sex, and BMI category. Specifically, a 1-SD higher AHEI or MDS was associated with a ≈5% greater peak VO_2_ (1.2 ml/kg/min at the sample mean); a magnitude similar to that observed for taking ≈4000 more steps per day in our previous work [3]. Adherence to healthy dietary patterns was also associated with multi-dimensional fitness measures reflecting distinct physiologies including autonomic function (rest and exercise HR), central cardiac and pulmonary vascular function (V_E_/VCO_2_) [23], and oxygen uptake kinetics (VO_2_/work). By merging metabolite associations with dietary indices and CRF, we observed broad directional consistency between metabolite relations with favorable dietary quality and CRF, highlighted metabolites previously implicated in cardiometabolic disease, and identified novel metabolites that may link the shared biology of diet and fitness.

Prior studies have demonstrated associations of dietary components with fitness, but have primarily focused on specific populations (e.g., extremes of age) [6–10], estimated measures of fitness [5,6,11–14], or specific dietary components [6,10,12,13]. Our study adds to the literature by directly addressing some of the limitations of prior investigations on dietary patterns and fitness. First, the use of CPET enabled quantification of peak VO_2_ for objective assessment of fitness and ensured that an adequate (i.e., diagnostic) level of volitional effort was provided by all participants (via the RER). By using comprehensive CPET, we were also able to measure the association of diet with different physiological components of fitness. For example, we observed the expected relation of higher carbohydrate intake with resting RER [27], which served as a positive control and supported the accuracy of our dietary indices. The relations of dietary quality with measures of autonomic (HR responses) and central cardiac/pulmonary vascular function (VE/VCO_2_) with exercise may also indicate that dietary quality has broader salutary effects beyond fuel substrate utilization on various organ systems.

Second, our large community-based sample enabled assessment of the relations between dietary quality and fitness throughout a broad age range and how they varied across sex, age, and BMI. We observed consistent associations (i.e., lack of effect modification) across sex and BMI categories, but there was a significant interaction between age and dietary quality scores on the association with percent predicted peak VO_2_, suggesting a higher effect size in younger individuals. This finding is consistent with prior studies that have often demonstrated associations between dietary and fitness in younger cohorts [6,9,10].

Third, we captured dietary patterns with a widely used FFQ and two healthy dietary indices designed to reflect the best contemporary evidence for healthy eating [19,20]. Both the AHEI and MDS have been robustly associated with lower risk of cardiovascular and all-cause mortality and other health outcomes [19,20,34,35], and their components/scores are readily translatable to individuals or for use in intervention studies. Furthermore, the use of dietary patterns captures synergies between food sources and better reflects how individuals typically eat. We observed that as percentages of total energy consumed, increased carbohydrate intake and decreased fat intake were associated with lower peak VO_2_. At the extremes, a very low-carbohydrate or ketogenic diet has been popularized for enhancing aerobic performance based on the theory that ketones may be more optimal fuel sources; however, the data for ketogenic diet improving peak VO_2_ in athletes are mixed [36]. A low-fat (<30% total energy intake) diet has been historically recommended for population health [37], but studies have increasingly shown that the quality and source of dietary fats and carbohydrates matter, leading to a focus on dietary patterns in recent guidelines [38]. In contrast to our findings, another observational study showed that lower fat intake was associated with higher CRF, but men in the study with high fitness also consumed less SFA (10.0% vs. 11.8% of total energy intake) and MUFA (12.6% vs. 14.5%) and a similar amount of PUFA (7.4% vs. 7.4%) compared to those with low fitness [13]. This discordance highlights the importance of food sources when considering macronutrient intake.

These epidemiological observations highlight the need for further investigation of potential mechanisms linking diet and CRF. To begin to address this gap, we explored potential shared biological processes by leveraging metabolite profiling. We identified 24 metabolites that were significantly associated with peak VO_2_, AHEI, and MDS after adjustment for physical activity and standard CVD risk factors. A particular strength of metabolomic interrogation in dietary research is that some metabolites may provide objective measures of the consumption of specific food items or the host-diet interaction. Indeed, several of our findings have been identified by prior studies to be associated with healthy dietary patterns [24,25,39–43]. Among our significant metabolites, those with previously reported positive associations with AHEI and MDS included lipid species such as C22:6 LPC, C38:7 PC plasmalogen, and C38:7 and C40:7 PE plasmalogens, which were all positively associated with greater fish intake. This finding is biochemically consistent with DHA being the fatty acid side chain in C22:6 LPC. Although our metabolomics platform did not precisely determine the fatty acid side chain composition of C38:7 PC plasmalogen, and C38:7 and C40:7 PE plasmalogens, DHA is likely one of their side chains based on their most plausible molecular structures. Omega-3 fatty acids are known to have favorable health benefits, and plasmalogens are hypothesized to have antioxidant properties and are inversely correlated with metabolic diseases [31]. It remains unclear whether these metabolites have benefits on CRF themselves versus being markers of omega-3 fatty acid intake. In small placebo-controlled pilot studies of athletes and sedentary adults, supplementation with DHA and EHA improved submaximal exercise HR, submaximal VO_2_, and/or peak VO_2_ [44–46], and in rat models, dietary omega-3 fatty acids reduced myocardial oxygen consumption while maintaining external work [47]. Potential mechanisms include increased altered mitochondrial membrane composition and mitochondrial biogenesis [48,49].

Metabolites with known inverse associations with AHEI and MDS included C6 and C7 carnitines, C18:1 sphingomyelin (SM), and C16:0 ceramide, which were also positively associated with intake of red meat, trans fatty acid, and SFA. Medium chain carnitines such as C6 and C7 carnitines are positively associated with obesity, type 2 diabetes, and CVD [28,50], and shorter chain sphingomyelins are positively associated with incident diabetes [51]. Ceramides have been recently identified as markers of incident CVD [29]; in mouse models, elevated C16:0 ceramide led to insulin resistance, and decreased C16:0 ceramide protected against diet-induced obesity [52]. Consequently, these metabolites are possibly linked to poor CRF through their relation with insulin resistance, which has been hypothesized to induce mitochondrial dysfunction [53] and may contribute to exercise intolerance.

In addition to the above known diet-related metabolites, we identified several metabolites displaying novel associations with dietary patterns and peak VO_2_. One notable example is N-monomethyl-arginine (NMMA), which inhibits nitric oxide synthase and causes increased vascular resistance [54]. Given this mechanism, its positive association with peak relative VO_2_ was surprising and may reflect improved autoregulation of blood pressure in those with higher CRF whereas those with chronically increased vascular resistance may have suppressed NMMA levels. In a randomized cross-over study of 10 healthy volunteers, the administration of L-NMMA increased resting blood pressure but did not change peripheral vascular resistance or cardiac output during or after exercise [55], and in a population study of African Americans, higher NMMA was associated with decreased risk of incident coronary artery disease [56]. Also related to the nitric oxide pathway, we observed significant positive associations of AHEI, MDS, and peak relative VO_2_ with N-acetyl-ornithine, which is an intermediate metabolite for arginine synthesis.

Overall, our analysis of metabolite associations with CRF and diet also implicated several metabolites with previously reported associations with cardiometabolic diseases. For example, higher cinnamoylglycine was associated with healthy dietary patterns and better CRF and was reported to be associated with decreased diabetes risk and improved insulin resistance [57]. By contrast, higher DMGV was associated with poorer dietary patterns and lower CRF and has been linked to poor metabolic health [58]. Given that CRF reflects underlying metabolic health, further studies are needed to determine whether the above metabolites are merely markers of dysmetabolism or whether they may have functional roles during exercise.

Our study provides a comprehensive assessment of the cross-sectional association between dietary quality and CRF in a large community-dwelling sample and uses shared associations with the circulating metabolome to infer potential biological mechanisms. Nevertheless, there are several limitations of the current investigation that warrant consideration. Despite adjustment for several potential confounders, we cannot exclude the possibility of residual confounding in this cross-sectional analysis. Future interventional studies are necessary to clarify whether the associations between diet and CRF are indeed causal. Moreover, how dietary choices at different time intervals prior to exercise (e.g., immediately before, in the days before, or in the weeks/months before) might influence CRF remains uncertain and would require prospective trials. Dietary quality assessment relied on self-recall and therefore may have limitations in regards to validity and reproducibility. While we do have several positive controls in our study that attest to the accuracy of the dietary information collected (e.g., peak RER and carbohydrate association, individual metabolites reflecting known food sources), the possibility for the measurement error of dietary quality cannot be excluded. Finally, despite the inclusion of the multi-ethnic Omni cohort, our study sample mostly consisted of White individuals eating American-style diets; the generalizability to other racial and ethnic groups is, therefore, unknown.

In conclusion, we showed in a large, community-dwelling cohort that a healthy diet is strongly associated with CRF and other physiologically relevant exercise responses. We identified several metabolites that shared associations with healthy patterns of eating and peak VO_2_, and many of these metabolites are known markers of cardiometabolic health. These findings provide a foundation for future work investigating whether dietary interventions that improve CRF may also impact long-term metabolic health.

## Data Availability

All data produced in the present study are available upon reasonable request to the Framingham Heart Study and the NIH's Database of Phenotypes and Genotypes.

## Author Contributions

M.Y.M., P.G., M.E.W., V.L.M., R.S.V., R.V.S., G.D.L., and M.N. designed research. M.E.W., P.M., V.X., R.S.V., R.V.S., G.D.L., and M.N. conducted research. M.Y.M., P.G., M.E.W., V.L.M., M.G.L., R.S.V., R.V.S., G.D.L., and M.N. analyzed data. M.Y.M., P.G., and M.N. wrote paper. M.N. had primary responsibility for final content. All authors have read and approved the final manuscript.

