## Supplementary Material for "Association of Healthy Dietary Patterns and Cardiorespiratory Fitness in the Community"

**Table of Contents**

Supplementary Material	Page

1. Supplementary Figure 1	2
2. Supplementary Figure 2	3
3. Supplementary Table 1	4
4. Supplementary Table 2	5
5. Supplementary Table 3	6
6. Supplementary Table 4	7
7. Supplementary Table 5	8
8. Supplementary Table 6	10
9. Supplementary Table 7	11
10. Supplementary References	18

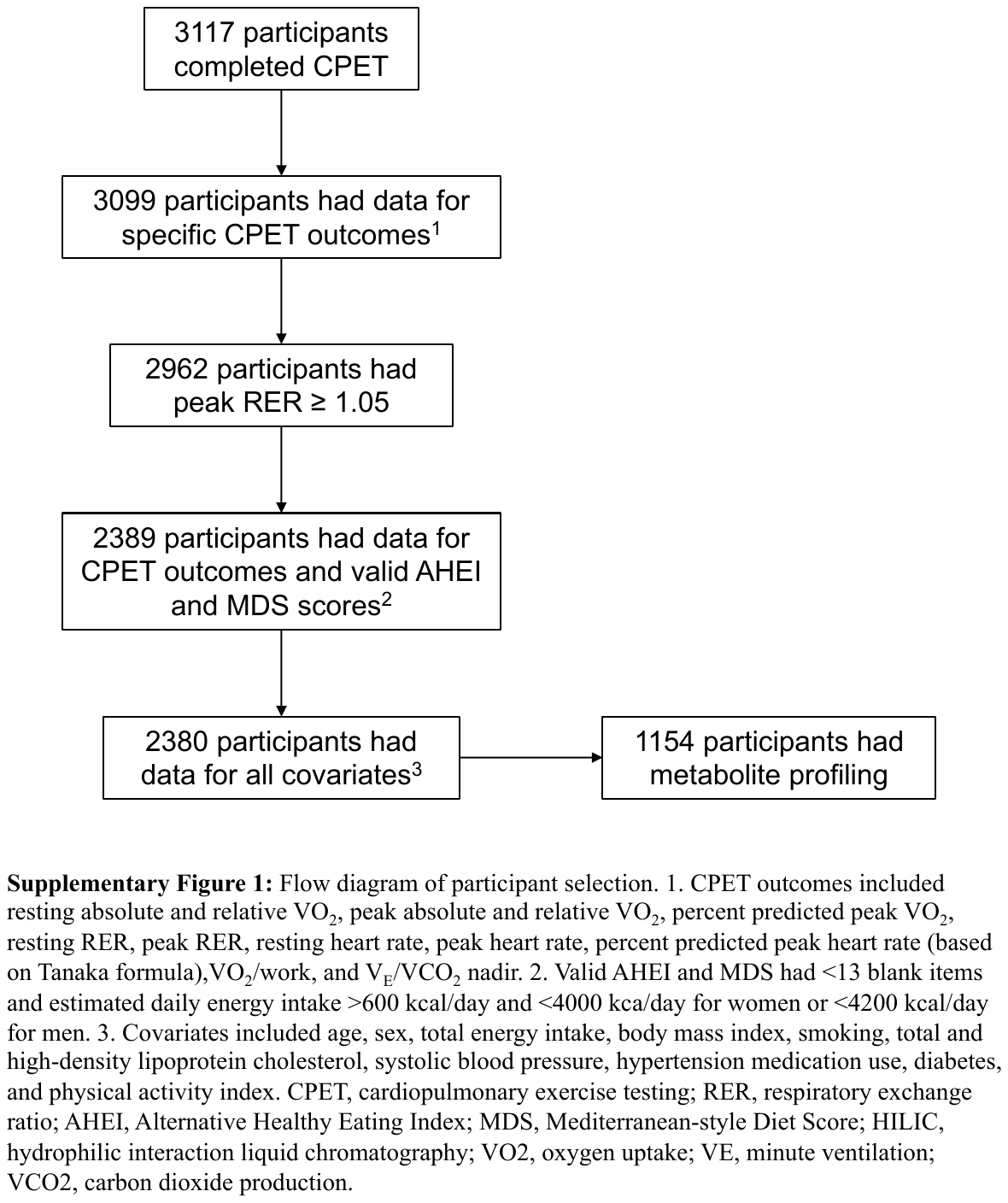

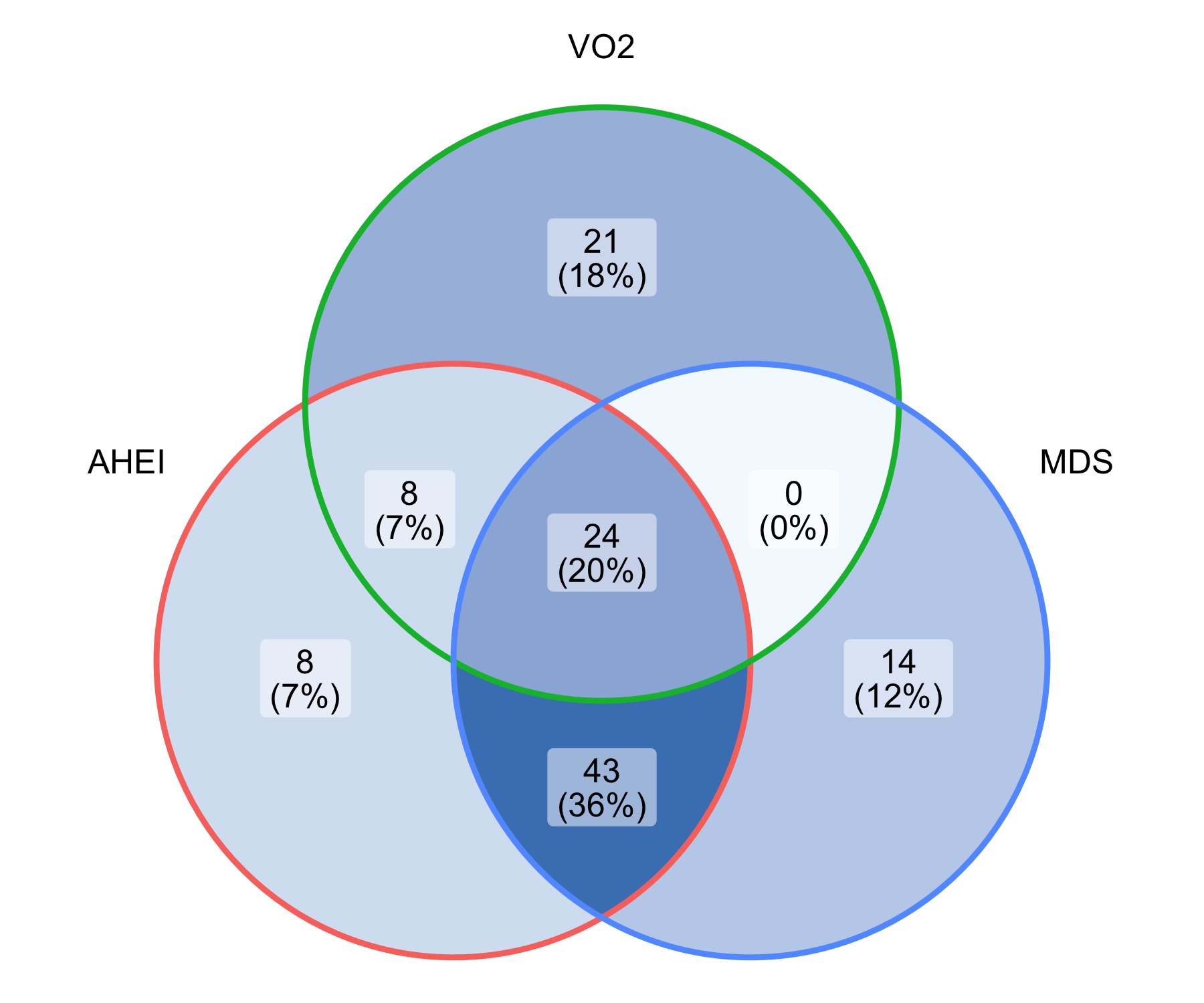

**Supplementary Figure 2:** Venn diagram of metabolite associations with Alternative Healthy Eating Index (AHEI), Mediterranean-style Diet Score (MDS), and peak relative oxygen uptake (VO_2_). Models were adjusted for age, sex, total energy intake, body mass index, smoking status, total cholesterol, high density lipoprotein cholesterol, systolic blood pressure, hypertension medication use, diabetes, and physical activity index.

**Supplementary Table 1:** Components of the Alternative Healthy Eating Index and Mediterranean-style Diet Score

|  | **Alternative Healthy Eating Index** | | | **Mediterranean-style Diet Score** | | |
| --- | --- | --- | --- | --- | --- | --- |
| **Component** | **Unit** | **Criteria for  minimum score (0)** | **Criteria for  maximum score (10)** | **Unit** | **Criteria for  minimum score (0)** | **Criteria for  maximum score (3)** |
| Vegetables | servings/day | 0 | ≥ 5 | servings/day | Lowest quartile | Highest quartile |
| Fruits | servings/day | 0 | ≥ 4 | servings/day | Lowest quartile | Highest quartile |
| Whole grains |  |  |  | servings/day | Lowest quartile | Highest quartile |
| Women | g/day |  | 75 |  |  |  |
| Men | g/day |  | 90 |  |  |  |
| Sugar-sweetened beverages  and fruit juice | servings/day | ≥ 1 | 0 |  |  |  |
| Nuts | servings/day | 0 | ≥ 1 | servings/day | Lowest quartile | Highest quartile |
| Legumes |  |  |  | servings/day | Lowest quartile | Highest quartile |
| Red meat | servings/day | ≥ 1.5 | 0 | servings/day | Highest quartile | Lowest quartile |
| Processed meat |  |  |  |  |  |  |
| Trans-fatty acids | % of energy | ≥ 4 | ≤ 0.5 |  |  |  |
| Long-chain (omega-3) fatty acids (EPA and DHA) | mg/day | 0 | 250 |  |  |  |
| Polyunsaturated fatty acids | % of energy | ≤ 2 | ≥ 10 |  |  |  |
| Sodium | mg/d | Highest decile | Lowest decile |  |  |  |
| Fish |  |  |  | servings/day | Lowest quartile | Highest quartile |
| Ratio of monounsaturated fatty acids to saturated fatty acids |  |  |  |  | Lowest quartile | Highest quartile |
| Alcohol |  |  |  |  |  |  |
| Women | drinks/day | ≥ 2.5 | 0.5 - 1.5 | g/day | 0 for all  other values | 1 if ≥ 5 g/day  and ≤ 15 g/day |
| Men | drinks/day | ≥ 3.5 | 0.5 - 2.0 | g/day | 0 for all  other values | 1 if ≥ 10 g/day  and ≤ 25 g/day |
| **Total** |  | **0** | **110** |  | **0** | **25** |

**Supplementary Table 2:** Cross sectional associations of dietary patterns and cardiopulmonary exercise testing variables after age, sex, and total caloric intake adjustment

|  | **Alternative Health Eating Index (N = 2380)** | | | **Mediterranean-style Diet Score (N = 2380)** | | |
| --- | --- | --- | --- | --- | --- | --- |
| **CPET Variables** | **Coefficient** | **SE** | **p-value** | **Coefficient** | **SE** | **p-value** |
| Resting Absolute VO_2_ | -0.074 | 0.016 | <0.0001 | -0.066 | 0.017 | 0.0001 |
| Resting Relative VO_2_ | 0.074 | 0.021 | 0.0004 | 0.092 | 0.021 | <0.0001 |
| Peak Absolute VO_2_ | 0.145 | 0.014 | <0.0001 | 0.124 | 0.014 | <0.0001 |
| Peak Relative VO_2_ | 0.272 | 0.017 | <0.0001 | 0.251 | 0.018 | <0.0001 |
| Percent Predicted Peak VO_2_ | 0.226 | 0.020 | <0.0001 | 0.197 | 0.021 | <0.0001 |
| Resting RER | -0.035 | 0.022 | 0.11 | -0.024 | 0.022 | 0.32 |
| Peak RER | 0.016 | 0.021 | 0.44 | 0.013 | 0.022 | 0.56 |
| Resting HR | -0.169 | 0.021 | <0.0001 | -0.154 | 0.022 | <0.0001 |
| Peak HR | 0.109 | 0.019 | <0.0001 | 0.122 | 0.019 | <0.0001 |
| Percent Predicted Peak HR | 0.120 | 0.021 | <0.0001 | 0.135 | 0.022 | <0.0001 |
| VO_2_/work | 0.147 | 0.020 | <0.0001 | 0.117 | 0.020 | <0.0001 |
| V_E_/VCO_2_ nadir | -0.112 | 0.020 | <0.0001 | -0.087 | 0.020 | <0.0001 |

Absolute and relative VO_2_ at rest and peak were log transformed. Coefficients represent one SD change in CPET variables per one SD increase in dietary score. Model adjusted for sex, age, and total caloric intake.

P-values were adjusted for false discovery rate of 5% for each predictor (diet) variable.

**Supplementary Table 3:** Cross sectional associations of dietary patterns and cardiopulmonary exercise testing variables with adjustment for objective physical activity measures

|  | **Alternative Health Eating Index (N = 1777)** | | | **Mediterranean-style Diet Score (N = 1777)** | | |
| --- | --- | --- | --- | --- | --- | --- |
| **CPET Variables** | **Coefficient** | **SE** | **p-value** | **Coefficient** | **SE** | **p-value** |
| Resting Absolute VO_2_ | 0.011 | 0.016 | 0.54 | 0.001 | 0.016 | 0.95 |
| Resting Relative VO_2_ | 0.012 | 0.022 | 0.58 | 0.005 | 0.022 | 0.91 |
| Peak Absolute VO_2_ | 0.126 | 0.016 | <0.0001 | 0.111 | 0.016 | <0.0001 |
| Peak Relative VO_2_ | 0.145 | 0.017 | <0.0001 | 0.131 | 0.017 | <0.0001 |
| Percent Predicted Peak VO_2_ | 0.181 | 0.024 | <0.0001 | 0.163 | 0.024 | <0.0001 |
| Resting RER | -0.023 | 0.027 | 0.47 | -0.018 | 0.028 | 0.63 |
| Peak RER | -0.025 | 0.025 | 0.43 | -0.022 | 0.026 | 0.52 |
| Resting HR | -0.114 | 0.024 | <0.0001 | -0.105 | 0.025 | 0.0001 |
| Peak HR | 0.035 | 0.021 | 0.16 | 0.061 | 0.022 | 0.0083 |
| Percent Predicted Peak HR | 0.038 | 0.024 | 0.16 | 0.068 | 0.024 | 0.0083 |
| VO_2_/work | 0.122 | 0.024 | <0.0001 | 0.090 | 0.024 | 0.0006 |
| V_E_/VCO_2_ | -0.095 | 0.023 | 0.0001 | -0.069 | 0.024 | 0.0083 |

Absolute and relative VO_2_ at rest and peak were log transformed. Coefficients represent one SD change in CPET variables per one SD increase in dietary score. Model adjusted for sex, age, total caloric intake, BMI, smoking status, cholesterol, HDL, systolic blood pressure, hypertension medication use, diabetes, sedentary time, and moderate-vigorous physical activity time. P-values were adjusted for false discovery rate of 5% for each predictor (diet) variable.

**Supplementary Table 4:** Cross sectional associations of dietary patterns and peak oxygen uptake and resting heart rate

|  | **Alternative Health Eating Index (N = 2380)** | | | | | | **Mediterranean-style Diet Score (N = 2380)** | | | | | |
| --- | --- | --- | --- | --- | --- | --- | --- | --- | --- | --- | --- | --- |
| **CPET Variables** | **Model 1** | | | **Model 2** | | | **Model 1** | | | **Model 2** | | |
|  | **Coefficient** | **SE** | **FDR p** | **Coefficient** | **SE** | **FDR p** | **Coefficient** | **SE** | **FDR p** | **Coefficient** | **SE** | **FDR p** |
| Resting Absolute VO_2_ | -0.032 | 0.015 | 0.039 | 0.028 | 0.013 | 0.04 | -0.028 | 0.016 | 0.079 | 0.036 | 0.014 | 0.011 |
| Resting Relative VO_2_ | 0.096 | 0.021 | <0.0001 | 0.033 | 0.018 | 0.072 | 0.112 | 0.021 | <0.0001 | 0.045 | 0.019 | 0.015 |
| Peak Absolute VO_2_ | 0.133 | 0.014 | <0.0001 | 0.136 | 0.013 | <0.0001 | 0.113 | 0.014 | <0.0001 | 0.117 | 0.014 | <0.0001 |
| Peak Relative VO_2_ | 0.236 | 0.017 | <0.0001 | 0.155 | 0.014 | <0.0001 | 0.217 | 0.017 | <0.0001 | 0.133 | 0.015 | <0.0001 |
| Percent Predicted Peak VO_2_ | 0.201 | 0.020 | <0.0001 | 0.192 | 0.020 | <0.0001 | 0.172 | 0.021 | <0.0001 | 0.164 | 0.021 | <0.0001 |
| Resting HR | -0.078 | 0.021 | 0.0002 | -0.080 | 0.021 | 0.0002 | -0.067 | 0.022 | 0.0021 | -0.069 | 0.021 | 0.0021 |

Absolute and relative VO_2_ at rest and peak were log transformed. Coefficients represent one SD change in CPET variables per one SD increase in dietary score. Model 1 adjusted for sex, age, and total caloric intake. Model 2 adjusted for sex, age, total caloric intake, BMI, smoking status, cholesterol, HDL, systolic blood pressure, hypertension medication use, diabetes, and physical activity index. VO_2_ variables were additionally adjusted for resting heart rate, and resting heart rate was adjusted for peak relative VO_2_ in both models. P-values were adjusted for false discovery rate of 5% for each predictor (diet) variable.

**Supplementary Table 5:** Regression coefficients for interaction of age, sex, and BMI on the associations of dietary patterns and CPET variables

| **CPET Variable** | **Diet** | **Subgroup** | **Coefficient** | **SE** | **Interaction p** | **FDR p** |
| --- | --- | --- | --- | --- | --- | --- |
| Peak Relative VO_2_ | AHEI | Male | 0.175 | 0.019 | - | - |
|  |  | Female | 0.165 | 0.020 | 0.70 | 0.94 |
|  | AHEI | Age <54 years | 0.169 | 0.019 | - | - |
|  |  | Age ≥54 years | 0.147 | 0.021 | 0.43 | 0.94 |
|  | AHEI | Normal | 0.173 | 0.024 | - | - |
|  |  | Overweight | 0.170 | 0.023 | 0.94 | 0.94 |
|  |  | Obese | 0.188 | 0.026 | 0.68 | 0.94 |
|  | MDS | Male | 0.162 | 0.020 | - | - |
|  |  | Female | 0.134 | 0.020 | 0.29 | 0.64 |
|  | MDS | Age <54 years | 0.147 | 0.020 | - | - |
|  |  | Age ≥54 years | 0.120 | 0.021 | 0.32 | 0.64 |
|  | MDS | Normal | 0.166 | 0.025 | -­ | - |
|  |  | Overweight | 0.151 | 0.024 | 0.65 | 0.74 |
|  |  | Obese | 0.154 | 0.026 | 0.74 | 0.74 |
| Resting Relative VO_2_ | AHEI | Male | 0.040 | 0.025 |  |  |
|  |  | Female | -0.033 | 0.026 | 0.04 | 0.06 |
|  | AHEI | Age <54 years | 0.001 | 0.024 |  |  |
|  |  | Age ≥54 years | -0.009 | 0.027 | 0.79 | 0.79 |
|  | AHEI | Normal | -0.053 | 0.031 |  |  |
|  |  | Overweight | 0.037 | 0.030 | 0.03 | 0.06 |
|  |  | Obese | 0.044 | 0.033 | 0.03 | 0.06 |
|  | MDS | Male | 0.047 | 0.026 |  |  |
|  |  | Female | -0.006 | 0.026 | 0.13 | 0.17 |
|  | MDS | Age <54 years | 0.021 | 0.025 |  |  |
|  |  | Age ≥54 years | -0.001 | 0.027 | 0.53 | 0.53 |
|  | MDS | Normal | -0.036 | 0.032 | ref | ref |
|  |  | Overweight | 0.052 | 0.030 | 0.04 | 0.07 |
|  |  | Obese | 0.064 | 0.033 | 0.02 | 0.07 |
| Percent Predicted Peak VO_2_ | AHEI | Male | 0.225 | 0.028 | - | - |
|  |  | Female | 0.198 | 0.029 | 0.49 | 0.56 |
|  | AHEI | Age <54 years | 0.291 | 0.026 | - | - |
|  |  | Age ≥54 years | 0.124 | 0.029 | <0.0001 | <0.0001 |
|  | AHEI | Normal | 0.181 | 0.034 | - | - |
|  |  | Overweight | 0.208 | 0.032 | 0.56 | 0.56 |
|  |  | Obese | 0.253 | 0.036 | 0.14 | 0.29 |
|  | MDS | Male | 0.214 | 0.028 | - | - |
|  |  | Female | 0.153 | 0.028 | 0.11 | 0.23 |
|  | MDS | Age <54 years | 0.249 | 0.027 | - | - |
|  |  | Age ≥54 years | 0.124 | 0.029 | 0.0009 | 0.004 |
|  | MDS | Normal | 0.178 | 0.034 | - | - |
|  |  | Overweight | 0.184 | 0.033 | 0.91 | 0.91 |
|  |  | Obese | 0.187 | 0.036 | 0.87 | 0.91 |

CPET, cardiopulmonary exercise test; FDR, false discovery rate; VO_2_, oxygen uptake; AHEI, Alternative Healthy Eating Index; MDS, Mediterranean-style Diet Score; BMI, body mass index. BMI categories defined as normal (BMI <25 kg/m^2^), overweight, (BMI 25-30 kg/m^2^), and obese (BMI ≥30 kg/m^2^). Models with sex*AHEI or sex*MDS interaction terms were additionally adjusted for age, BMI, total energy intake, smoking status, total cholesterol, high-density lipoprotein cholesterol, systolic blood pressure, hypertension medication use, diabetes, and physical activity index. Models with median-split age*AHEI or median-split age*MDS interaction terms were additionally adjusted for sex, BMI, total energy intake, smoking status, total cholesterol, high-density lipoprotein cholesterol, systolic blood pressure, hypertension medication use, diabetes, and physical activity index. Models with BMI category*AHEI or BMI category*MDS interaction terms were additionally adjusted for age, sex, total energy intake, smoking status, total cholesterol, high-density lipoprotein cholesterol, systolic blood pressure, hypertension medication use, diabetes, and physical activity index. FDR applied across CPET variables (peak relative VO_2_, resting relative VO_2_, percent predicted peak VO_2_) and diet indices (AHEI and MDS).

**Supplementary Table 6:** Cross sectional associations of macronutrient intake and cardiopulmonary exercise testing variables

|  | **Percent Energy Intake from Carbohydrate (N = 2380)** | | | | | | **Percent Energy Intake from Fat (N = 2380)** | | | | | | **Percent Energy Intake from Protein (N = 2380)** | | | | | |
| --- | --- | --- | --- | --- | --- | --- | --- | --- | --- | --- | --- | --- | --- | --- | --- | --- | --- | --- |
| **CPET Variables** | **Model 1** | | | **Model 2** | | | **Model 1** | | | **Model 2** | | | **Model 1** | | | **Model 2** | | |
|  | **Coefficient** | **SE** | **FDR p** | **Coefficient** | **SE** | **FDR p** | **Coefficient** | **SE** | **FDR p** | **Coefficient** | **SE** | **FDR p** | **Coefficient** | **SE** | **FDR p** | **Coefficient** | **SE** | **FDR p** |
| Resting Absolute VO_2_ | -0.025 | 0.005 | <0.0001 | -0.011 | 0.005 | 0.024 | 0.010 | 0.005 | 0.1296 | 0.001 | 0.004 | 0.92 | 0.019 | 0.006 | 0.0044 | -0.004 | 0.005 | 0.6460 |
| Resting Relative VO_2_ | 0.001 | 0.007 | 0.91 | -0.007 | 0.006 | 0.26 | -0.018 | 0.007 | 0.0265 | -0.003 | 0.006 | 0.85 | -0.032 | 0.007 | 0.0001 | -0.007 | 0.006 | 0.6460 |
| Peak Absolute VO_2_ | -0.027 | 0.005 | <0.0001 | -0.027 | 0.004 | <0.0001 | 0.023 | 0.005 | <0.0001 | 0.021 | 0.004 | <0.0001 | 0.011 | 0.005 | 0.049 | 0.004 | 0.005 | 0.6460 |
| Peak Relative VO_2_ | -0.010 | 0.006 | 0.16 | -0.027 | 0.005 | <0.0001 | 0.008 | 0.006 | 0.3746 | 0.021 | 0.005 | <0.0001 | -0.020 | 0.006 | 0.0044 | 0.004 | 0.005 | 0.6460 |
| Percent Predicted Peak VO_2_ | -0.036 | 0.007 | <0.0001 | -0.040 | 0.007 | <0.0001 | 0.031 | 0.007 | <0.0001 | 0.030 | 0.007 | <0.0001 | 0.009 | 0.007 | 0.26 | 0.003 | 0.007 | 0.6921 |
| Resting RER | 0.031 | 0.007 | <0.0001 | 0.031 | 0.007 | <0.0001 | -0.008 | 0.007 | 0.4977 | -0.009 | 0.007 | 0.39 | -0.020 | 0.007 | 0.023 | -0.025 | 0.007 | 0.0104 |
| Peak RER | 0.026 | 0.007 | 0.0006 | 0.015 | 0.007 | 0.040 | -0.006 | 0.007 | 0.6115 | 0.002 | 0.007 | 0.92 | -0.007 | 0.007 | 0.40 | 0.004 | 0.007 | 0.6479 |
| Resting HR | -0.010 | 0.007 | 0.18 | -0.003 | 0.007 | 0.72 | -0.003 | 0.007 | 0.8197 | -0.007 | 0.007 | 0.45 | -0.007 | 0.007 | 0.40 | -0.018 | 0.007 | 0.0570 |
| Peak HR | -0.009 | 0.006 | 0.18 | -0.019 | 0.006 | 0.0027 | 0.002 | 0.006 | 0.8197 | 0.010 | 0.006 | 0.19 | -0.012 | 0.006 | 0.088 | -0.004 | 0.006 | 0.6460 |
| Percent Predicted Peak HR | -0.010 | 0.007 | 0.18 | -0.021 | 0.007 | 0.0027 | 0.002 | 0.007 | 0.8197 | 0.011 | 0.007 | 0.19 | -0.014 | 0.007 | 0.088 | -0.005 | 0.007 | 0.6460 |
| VO_2_/work | -0.032 | 0.007 | <0.0001 | -0.033 | 0.007 | <0.0001 | 0.035 | 0.007 | <0.0001 | 0.033 | 0.007 | <0.0001 | 0.005 | 0.007 | 0.47 | 0.003 | 0.007 | 0.6921 |
| V_E_/VCO_2_ | -0.008 | 0.007 | 0.24 | -0.010 | 0.007 | 0.18 | <0.001 | 0.007 | 0.9721 | <0.001 | 0.007 | 0.98 | -0.015 | 0.007 | 0.056 | -0.013 | 0.007 | 0.1878 |

Absolute and relative VO_2_ at rest and peak were log transformed. Coefficients represent one SD change in CPET variables per one decile increase in percent energy intake. Model 1 adjusted for sex, age, and total caloric intake. Model 2 adjusted for sex, age, total caloric intake, BMI, smoking status, cholesterol, HDL, systolic blood pressure, hypertension medication use, diabetes, and physical activity index. P-values were adjusted for false discovery rate of 5% within each column.

**Supplementary Table 7:** Metabolite associations with dietary patterns and CPET variables

|  | **Alternative Healthy Eating Index (N = 1154)** | | | **Mediterranean-style Diet Score (N = 1154)** | | | **Peak Relative VO_2_ (N = 1154)** | | |
| --- | --- | --- | --- | --- | --- | --- | --- | --- | --- |
| **Metabolite** | **Coefficient** | **SE** | **FDR** | **Coefficient** | **SE** | **FDR** | **Coefficient** | **SE** | **FDR** |
| 1-methyladenosine | -0.022 | 0.029 | 0.584 | -0.084 | 0.029 | 0.014 | -0.018 | 0.040 | 0.794 |
| 1-methylguanine | -0.005 | 0.031 | 0.893 | -0.037 | 0.032 | 0.384 | 0.053 | 0.044 | 0.402 |
| 1-methylguanosine | -0.061 | 0.030 | 0.095 | -0.083 | 0.031 | 0.020 | 0.069 | 0.042 | 0.229 |
| 1-methylnicotinamide | 0.091 | 0.031 | 0.012 | 0.024 | 0.032 | 0.585 | 0.139 | 0.044 | 0.010 |
| 1,7-dimethyluric acid | 0.059 | 0.031 | 0.115 | 0.030 | 0.032 | 0.497 | 0.067 | 0.043 | 0.262 |
| 2-aminooctanoate | 0.077 | 0.031 | 0.037 | 0.098 | 0.032 | 0.008 | 0.091 | 0.044 | 0.113 |
| 2-methylguanosine | -0.065 | 0.030 | 0.064 | -0.051 | 0.030 | 0.173 | -0.095 | 0.042 | 0.074 |
| 3-hydroxyanthranilic acid | -0.019 | 0.028 | 0.629 | -0.057 | 0.029 | 0.105 | 0.005 | 0.040 | 0.926 |
| 3-methylhistidine | 0.087 | 0.031 | 0.016 | 0.070 | 0.032 | 0.068 | 0.139 | 0.044 | 0.010 |
| 3-methylxanthine | 0.036 | 0.031 | 0.374 | -0.007 | 0.032 | 0.899 | 0.140 | 0.044 | 0.010 |
| 4-acetamidobutanoate | 0.013 | 0.029 | 0.730 | 0.017 | 0.030 | 0.696 | 0.019 | 0.041 | 0.794 |
| 4-hydroxyhippurate | 0.097 | 0.032 | 0.008 | 0.092 | 0.033 | 0.014 | 0.052 | 0.045 | 0.415 |
| 5-acetylamino-6-amino-3-methyluracil | 0.057 | 0.031 | 0.124 | 0.026 | 0.031 | 0.555 | 0.062 | 0.043 | 0.297 |
| 5-hydroxymethyl-4-methyluracil | -0.022 | 0.032 | 0.615 | 0.007 | 0.032 | 0.899 | -0.042 | 0.044 | 0.528 |
| 5-hydroxytryptophol | 0.119 | 0.031 | 0.001 | 0.086 | 0.032 | 0.021 | 0.003 | 0.044 | 0.954 |
| 7-methylguanine | -0.023 | 0.031 | 0.593 | -0.019 | 0.032 | 0.681 | 0.009 | 0.043 | 0.914 |
| 7-methylxanthine | -0.004 | 0.032 | 0.920 | -0.051 | 0.033 | 0.205 | 0.117 | 0.045 | 0.037 |
| acetylgalactosamine | -0.013 | 0.031 | 0.739 | -0.011 | 0.031 | 0.830 | -0.080 | 0.043 | 0.154 |
| alanine | -0.015 | 0.029 | 0.698 | -0.028 | 0.030 | 0.499 | 0.046 | 0.041 | 0.430 |
| allantoin | -0.032 | 0.029 | 0.402 | -0.025 | 0.030 | 0.556 | -0.019 | 0.041 | 0.794 |
| alpha glycerophosphocholine | -0.091 | 0.030 | 0.009 | -0.107 | 0.031 | 0.002 | -0.006 | 0.042 | 0.926 |
| aminoisobutyric acid | 0.052 | 0.031 | 0.169 | 0.079 | 0.032 | 0.035 | 0.071 | 0.044 | 0.229 |
| arginine | 0.051 | 0.031 | 0.183 | 0.008 | 0.032 | 0.892 | 0.047 | 0.044 | 0.453 |
| asparagine | 0.094 | 0.031 | 0.008 | 0.094 | 0.032 | 0.010 | 0.156 | 0.043 | 0.004 |
| betaine | 0.093 | 0.029 | 0.006 | 0.102 | 0.030 | 0.003 | 0.072 | 0.041 | 0.190 |
| bilirubin | 0.044 | 0.030 | 0.255 | 0.023 | 0.031 | 0.596 | 0.164 | 0.042 | 0.002 |
| biliverdin | 0.005 | 0.030 | 0.893 | -0.016 | 0.031 | 0.704 | 0.136 | 0.042 | 0.009 |
| butyrobetaine | -0.061 | 0.027 | 0.052 | -0.071 | 0.027 | 0.024 | 0.087 | 0.037 | 0.068 |
| C10 carnitine | -0.081 | 0.031 | 0.028 | -0.092 | 0.032 | 0.013 | -0.067 | 0.044 | 0.267 |
| C10:2 carnitine | 0.009 | 0.031 | 0.817 | -0.016 | 0.032 | 0.714 | -0.024 | 0.044 | 0.751 |
| C12 carnitine | -0.135 | 0.031 | 0.0001 | -0.133 | 0.032 | 0.0002 | -0.033 | 0.044 | 0.631 |
| C12:1 carnitine | -0.106 | 0.031 | 0.003 | -0.092 | 0.032 | 0.013 | -0.080 | 0.043 | 0.167 |
| C14 carnitine | -0.174 | 0.030 | <0.0001 | -0.179 | 0.031 | <0.0001 | -0.063 | 0.043 | 0.285 |
| C14:0 LPC | -0.040 | 0.030 | 0.300 | -0.062 | 0.031 | 0.096 | 0.049 | 0.042 | 0.417 |
| C14:0 SM | -0.041 | 0.024 | 0.148 | -0.090 | 0.024 | 0.001 | 0.041 | 0.033 | 0.396 |
| C14:1 carnitine | -0.054 | 0.031 | 0.146 | -0.025 | 0.031 | 0.557 | -0.046 | 0.043 | 0.448 |
| C14:2 carnitine | -0.082 | 0.031 | 0.023 | -0.058 | 0.032 | 0.134 | -0.066 | 0.043 | 0.263 |
| C16 carnitine | -0.124 | 0.028 | 0.0001 | -0.130 | 0.029 | 0.0001 | -0.095 | 0.040 | 0.057 |
| C16:0 ceramide d18:1 | -0.126 | 0.028 | 0.0001 | -0.096 | 0.029 | 0.003 | -0.122 | 0.039 | 0.012 |
| C16:0 LPC | -0.003 | 0.028 | 0.915 | -0.016 | 0.028 | 0.691 | 0.063 | 0.039 | 0.234 |
| C16:0 LPE | 0.038 | 0.029 | 0.300 | 0.041 | 0.030 | 0.294 | 0.050 | 0.041 | 0.396 |
| C16:0 SM | -0.029 | 0.022 | 0.307 | -0.013 | 0.023 | 0.691 | 0.037 | 0.031 | 0.412 |
| C16:1 LPC | -0.133 | 0.030 | 0.0001 | -0.131 | 0.030 | 0.0001 | -0.066 | 0.042 | 0.254 |
| C16:1 LPC plasmalogen | -0.028 | 0.028 | 0.457 | -0.038 | 0.029 | 0.325 | -0.008 | 0.040 | 0.914 |
| C16:1 SM | -0.043 | 0.020 | 0.076 | -0.039 | 0.021 | 0.131 | -0.101 | 0.029 | 0.004 |
| C18 carnitine | -0.103 | 0.030 | 0.003 | -0.148 | 0.030 | <0.0001 | 0.048 | 0.042 | 0.418 |
| C18:0 LPC | -0.044 | 0.027 | 0.196 | -0.045 | 0.028 | 0.188 | 0.031 | 0.038 | 0.614 |
| C18:0 LPE | -0.032 | 0.028 | 0.380 | -0.030 | 0.029 | 0.435 | 0.012 | 0.039 | 0.878 |
| C18:0 LPE B | -0.080 | 0.030 | 0.025 | -0.111 | 0.031 | 0.002 | 0.007 | 0.043 | 0.920 |
| C18:0 SM | -0.138 | 0.026 | <0.0001 | -0.132 | 0.027 | <0.0001 | -0.081 | 0.037 | 0.095 |
| C18:1 carnitine | 0.037 | 0.030 | 0.336 | 0.074 | 0.031 | 0.042 | -0.008 | 0.042 | 0.914 |
| C18:1 LPC | 0.034 | 0.027 | 0.318 | 0.058 | 0.027 | 0.079 | 0.127 | 0.037 | 0.005 |
| C18:1 LPC plasmalogen | 0.066 | 0.029 | 0.052 | 0.100 | 0.030 | 0.003 | 0.035 | 0.041 | 0.577 |
| C18:1 LPE | -0.032 | 0.030 | 0.400 | <0.0001 | 0.031 | 0.999 | 0.042 | 0.042 | 0.487 |
| C18:1 SM | -0.132 | 0.024 | <0.0001 | -0.126 | 0.025 | <0.0001 | -0.141 | 0.034 | 0.001 |
| C18:2 carnitine | -0.015 | 0.028 | 0.695 | -0.002 | 0.029 | 0.970 | -0.095 | 0.039 | 0.057 |
| C18:2 LPC | -0.008 | 0.027 | 0.813 | -0.016 | 0.027 | 0.691 | 0.118 | 0.037 | 0.010 |
| C18:2 LPE | -0.116 | 0.030 | 0.001 | -0.109 | 0.031 | 0.002 | -0.019 | 0.042 | 0.794 |
| C18:3 LPC | 0.005 | 0.030 | 0.895 | -0.026 | 0.030 | 0.546 | 0.028 | 0.041 | 0.667 |
| C18:3 LPE | -0.032 | 0.030 | 0.402 | -0.031 | 0.031 | 0.459 | -0.012 | 0.042 | 0.891 |
| C2 carnitine | -0.021 | 0.030 | 0.618 | -0.008 | 0.031 | 0.886 | 0.008 | 0.042 | 0.914 |
| C20:0 LPE | -0.057 | 0.029 | 0.106 | -0.085 | 0.030 | 0.014 | 0.016 | 0.041 | 0.832 |
| C20:0 SM | 0.006 | 0.027 | 0.864 | -0.015 | 0.028 | 0.704 | 0.061 | 0.038 | 0.239 |
| C20:1 LPC | 0.168 | 0.026 | <0.0001 | 0.210 | 0.026 | <0.0001 | 0.154 | 0.036 | 0.001 |
| C20:1 LPE | -0.169 | 0.030 | <0.0001 | -0.185 | 0.030 | <0.0001 | -0.059 | 0.042 | 0.314 |
| C20:3 LPC | -0.096 | 0.029 | 0.004 | -0.096 | 0.030 | 0.005 | 0.051 | 0.041 | 0.396 |
| C20:4 LPC | -0.075 | 0.029 | 0.027 | -0.070 | 0.030 | 0.047 | 0.005 | 0.041 | 0.926 |
| C20:4 LPE | -0.166 | 0.030 | <0.0001 | -0.154 | 0.031 | <0.0001 | -0.102 | 0.043 | 0.057 |
| C20:5 LPC | 0.101 | 0.028 | 0.002 | 0.057 | 0.029 | 0.100 | 0.143 | 0.039 | 0.004 |
| C22:0 SM | 0.008 | 0.028 | 0.817 | 0.005 | 0.029 | 0.913 | 0.107 | 0.040 | 0.032 |
| C22:4 LPC | -0.229 | 0.029 | <0.0001 | -0.206 | 0.029 | <0.0001 | -0.033 | 0.041 | 0.614 |
| C22:5 LPC | -0.136 | 0.029 | <0.0001 | -0.146 | 0.029 | <0.0001 | 0.012 | 0.040 | 0.878 |
| C22:6 LPC | 0.205 | 0.029 | <0.0001 | 0.179 | 0.030 | <0.0001 | 0.172 | 0.041 | 0.001 |
| C22:6 LPE | 0.196 | 0.029 | <0.0001 | 0.165 | 0.030 | <0.0001 | 0.058 | 0.042 | 0.314 |
| C24:0 LPC | 0.208 | 0.025 | <0.0001 | 0.210 | 0.025 | <0.0001 | 0.133 | 0.036 | 0.003 |
| C24:1 ceramide d18:1 | -0.040 | 0.029 | 0.272 | -0.023 | 0.030 | 0.585 | -0.070 | 0.041 | 0.204 |
| C26 carnitine | 0.070 | 0.030 | 0.045 | 0.052 | 0.030 | 0.161 | 0.086 | 0.041 | 0.113 |
| C3 carnitine | -0.005 | 0.027 | 0.893 | -0.007 | 0.028 | 0.892 | -0.079 | 0.038 | 0.113 |
| C3:DC CH3 carnitine | -0.077 | 0.028 | 0.021 | -0.074 | 0.029 | 0.031 | -0.070 | 0.040 | 0.190 |
| C30:0 PC | -0.057 | 0.028 | 0.089 | -0.066 | 0.029 | 0.050 | 0.061 | 0.039 | 0.257 |
| C32:2 PC | -0.043 | 0.029 | 0.236 | -0.052 | 0.029 | 0.148 | -0.024 | 0.040 | 0.717 |
| C34:0 PE | -0.099 | 0.030 | 0.004 | -0.147 | 0.030 | <0.0001 | 0.031 | 0.042 | 0.642 |
| C34:1 DAG or TAG fragment | -0.080 | 0.026 | 0.008 | -0.072 | 0.026 | 0.019 | -0.134 | 0.036 | 0.003 |
| C34:2 DAG or TAG fragment | -0.042 | 0.027 | 0.214 | -0.057 | 0.028 | 0.090 | -0.124 | 0.038 | 0.009 |
| C34:2 PE | -0.052 | 0.030 | 0.152 | -0.031 | 0.030 | 0.459 | -0.043 | 0.042 | 0.472 |
| C34:2 PE plasmalogen | 0.013 | 0.029 | 0.722 | -0.020 | 0.029 | 0.619 | 0.100 | 0.040 | 0.050 |
| C34:3 PC | -0.065 | 0.028 | 0.050 | -0.067 | 0.029 | 0.047 | -0.094 | 0.039 | 0.057 |
| C34:3 PC plasmalogen | -0.015 | 0.026 | 0.677 | -0.045 | 0.027 | 0.173 | 0.105 | 0.037 | 0.022 |
| C34:3 PE plasmalogen | -0.042 | 0.031 | 0.276 | -0.092 | 0.031 | 0.011 | 0.053 | 0.043 | 0.396 |
| C34:4 PC | -0.043 | 0.030 | 0.263 | -0.054 | 0.031 | 0.153 | -0.048 | 0.042 | 0.418 |
| C36:2 PC | -0.058 | 0.029 | 0.100 | -0.039 | 0.030 | 0.316 | 0.008 | 0.041 | 0.914 |
| C36:2 PE | -0.019 | 0.029 | 0.629 | -0.003 | 0.030 | 0.964 | -0.044 | 0.041 | 0.448 |
| C36:2 PS plasmalogen | 0.024 | 0.026 | 0.483 | 0.028 | 0.026 | 0.430 | -0.027 | 0.036 | 0.635 |
| C36:3 PE plasmalogen | -0.070 | 0.029 | 0.044 | -0.113 | 0.030 | 0.001 | 0.063 | 0.041 | 0.266 |
| C36:3 PS plasmalogen | -0.079 | 0.029 | 0.018 | -0.091 | 0.029 | 0.007 | -0.027 | 0.040 | 0.680 |
| C36:4 PE | -0.039 | 0.030 | 0.307 | -0.006 | 0.031 | 0.899 | -0.053 | 0.042 | 0.396 |
| C36:5 PC plasmalogen | -0.087 | 0.029 | 0.008 | -0.099 | 0.029 | 0.003 | -0.039 | 0.040 | 0.514 |
| C36:5 PE plasmalogen | -0.015 | 0.030 | 0.715 | -0.041 | 0.031 | 0.316 | 0.003 | 0.043 | 0.954 |
| C38:4 PE | -0.081 | 0.029 | 0.016 | -0.050 | 0.030 | 0.173 | -0.105 | 0.041 | 0.043 |
| C38:5 PE plasmalogen | -0.163 | 0.030 | <0.0001 | -0.182 | 0.031 | <0.0001 | -0.083 | 0.043 | 0.138 |
| C38:6 PC plasmalogen | 0.110 | 0.026 | 0.0002 | 0.096 | 0.027 | 0.002 | 0.027 | 0.037 | 0.642 |
| C38:6 PE | 0.225 | 0.029 | <0.0001 | 0.225 | 0.030 | <0.0001 | 0.081 | 0.042 | 0.138 |
| C38:6 PE plasmalogen | 0.028 | 0.029 | 0.464 | -0.009 | 0.030 | 0.870 | 0.026 | 0.040 | 0.695 |
| C38:7 PC plasmalogen | 0.266 | 0.027 | <0.0001 | 0.221 | 0.028 | <0.0001 | 0.137 | 0.040 | 0.005 |
| C38:7 PE plasmalogen | 0.324 | 0.028 | <0.0001 | 0.274 | 0.029 | <0.0001 | 0.177 | 0.041 | 0.001 |
| C4 carnitine | 0.017 | 0.031 | 0.675 | -0.002 | 0.031 | 0.976 | 0.009 | 0.043 | 0.914 |
| C4:OH carnitine | 0.024 | 0.029 | 0.549 | 0.025 | 0.030 | 0.555 | 0.029 | 0.041 | 0.653 |
| C40 6 PE | 0.139 | 0.028 | <0.0001 | 0.137 | 0.029 | <0.0001 | 0.015 | 0.040 | 0.837 |
| C40:7 PE plasmalogen | 0.188 | 0.027 | <0.0001 | 0.136 | 0.028 | <0.0001 | 0.114 | 0.039 | 0.018 |
| C5 carnitine | -0.019 | 0.027 | 0.595 | -0.058 | 0.027 | 0.080 | 0.008 | 0.038 | 0.914 |
| C5 DC carnitine | -0.004 | 0.031 | 0.915 | -0.014 | 0.032 | 0.757 | 0.059 | 0.044 | 0.339 |
| C5:1 carnitine | 0.104 | 0.031 | 0.003 | 0.074 | 0.031 | 0.046 | 0.151 | 0.043 | 0.004 |
| C6 carnitine | -0.107 | 0.030 | 0.002 | -0.131 | 0.031 | 0.0001 | -0.150 | 0.042 | 0.004 |
| C7 carnitine | -0.195 | 0.030 | <0.0001 | -0.230 | 0.031 | <0.0001 | -0.113 | 0.043 | 0.037 |
| C8 carnitine | -0.085 | 0.031 | 0.020 | -0.109 | 0.032 | 0.003 | -0.087 | 0.044 | 0.137 |
| C9 carnitine | -0.049 | 0.032 | 0.214 | -0.088 | 0.032 | 0.019 | 0.024 | 0.044 | 0.753 |
| carnitine | -0.027 | 0.029 | 0.477 | -0.008 | 0.030 | 0.879 | -0.029 | 0.041 | 0.650 |
| choline | -0.018 | 0.028 | 0.632 | -0.052 | 0.028 | 0.132 | 0.088 | 0.038 | 0.074 |
| cinnamoylglycine | 0.188 | 0.029 | <0.0001 | 0.184 | 0.030 | <0.0001 | 0.182 | 0.041 | 0.001 |
| citrulline | -0.041 | 0.029 | 0.274 | -0.063 | 0.030 | 0.082 | 0.049 | 0.041 | 0.409 |
| cortisol | -0.001 | 0.032 | 0.989 | 0.016 | 0.032 | 0.723 | 0.034 | 0.044 | 0.631 |
| cortisone | 0.055 | 0.031 | 0.145 | 0.058 | 0.032 | 0.134 | 0.082 | 0.043 | 0.150 |
| creatine | -0.014 | 0.028 | 0.715 | -0.053 | 0.029 | 0.132 | -0.168 | 0.039 | 0.001 |
| creatinine | -0.015 | 0.026 | 0.666 | -0.020 | 0.027 | 0.585 | 0.108 | 0.036 | 0.017 |
| cyclohexylamine | -0.018 | 0.032 | 0.668 | -0.010 | 0.032 | 0.856 | -0.026 | 0.044 | 0.717 |
| cystine | 0.063 | 0.029 | 0.061 | 0.020 | 0.029 | 0.619 | -0.044 | 0.040 | 0.441 |
| cytosine | 0.071 | 0.031 | 0.052 | 0.063 | 0.032 | 0.102 | 0.105 | 0.043 | 0.057 |
| dimethylglycine | 0.019 | 0.029 | 0.629 | 0.027 | 0.029 | 0.500 | 0.043 | 0.040 | 0.453 |
| DMGV | -0.092 | 0.024 | 0.001 | -0.117 | 0.025 | <0.0001 | -0.132 | 0.034 | 0.002 |
| ectoine | 0.166 | 0.031 | <0.0001 | 0.143 | 0.032 | 0.0001 | 0.122 | 0.044 | 0.027 |
| GABA | 0.108 | 0.030 | 0.002 | 0.067 | 0.031 | 0.078 | 0.163 | 0.042 | 0.002 |
| glutamate | -0.029 | 0.023 | 0.324 | -0.024 | 0.023 | 0.450 | -0.092 | 0.032 | 0.021 |
| glutamine | 0.059 | 0.031 | 0.111 | 0.064 | 0.031 | 0.090 | 0.072 | 0.043 | 0.219 |
| glycine | 0.012 | 0.028 | 0.735 | -0.007 | 0.029 | 0.887 | -0.007 | 0.039 | 0.914 |
| glycocholate | -0.036 | 0.031 | 0.364 | -0.036 | 0.032 | 0.404 | -0.116 | 0.044 | 0.035 |
| glycodeoxycholate/  glycochenodeoxycholate | -0.058 | 0.031 | 0.126 | -0.053 | 0.032 | 0.173 | -0.174 | 0.044 | 0.002 |
| guanidinoacetic acid | -0.011 | 0.029 | 0.759 | -0.032 | 0.030 | 0.430 | 0.025 | 0.041 | 0.708 |
| histamine | 0.062 | 0.032 | 0.108 | -0.041 | 0.032 | 0.341 | 0.020 | 0.044 | 0.794 |
| histidine | 0.096 | 0.031 | 0.008 | 0.035 | 0.032 | 0.430 | 0.069 | 0.044 | 0.257 |
| homoarginine | 0.053 | 0.029 | 0.134 | 0.032 | 0.030 | 0.430 | -0.020 | 0.041 | 0.788 |
| homocitrulline | 0.075 | 0.031 | 0.038 | 0.057 | 0.032 | 0.135 | 0.134 | 0.043 | 0.012 |
| hydroxyproline | -0.054 | 0.031 | 0.149 | -0.113 | 0.032 | 0.002 | -0.001 | 0.044 | 0.980 |
| imidazole propionate | -0.072 | 0.028 | 0.029 | -0.064 | 0.029 | 0.060 | -0.048 | 0.039 | 0.402 |
| imidazoleacetic acid | 0.056 | 0.031 | 0.135 | 0.025 | 0.031 | 0.557 | 0.117 | 0.043 | 0.030 |
| isoleucine | 0.007 | 0.025 | 0.819 | 0.005 | 0.025 | 0.899 | 0.042 | 0.034 | 0.396 |
| kynurenic acid | 0.017 | 0.030 | 0.668 | -0.005 | 0.031 | 0.913 | 0.087 | 0.042 | 0.110 |
| L-glutamyl-L-lysine | 0.016 | 0.031 | 0.698 | -0.001 | 0.032 | 0.992 | -0.003 | 0.044 | 0.954 |
| leucine | 0.046 | 0.024 | 0.123 | 0.024 | 0.025 | 0.469 | 0.098 | 0.034 | 0.021 |
| linoleoyl ethanolamide | -0.071 | 0.032 | 0.061 | -0.078 | 0.033 | 0.045 | -0.052 | 0.045 | 0.418 |
| lysine | 0.042 | 0.031 | 0.285 | 0.002 | 0.032 | 0.979 | -0.009 | 0.043 | 0.914 |
| methionine | -0.036 | 0.029 | 0.324 | -0.025 | 0.030 | 0.555 | -0.008 | 0.041 | 0.914 |
| methionine sulfoxide | 0.088 | 0.031 | 0.016 | 0.039 | 0.032 | 0.361 | 0.040 | 0.044 | 0.540 |
| methylimidazoleacetic acid | 0.071 | 0.031 | 0.052 | 0.053 | 0.032 | 0.174 | 0.124 | 0.044 | 0.023 |
| myristoleate | -0.125 | 0.031 | 0.0004 | -0.131 | 0.032 | 0.0002 | -0.026 | 0.044 | 0.717 |
| N-acetylalanine | -0.028 | 0.031 | 0.483 | -0.028 | 0.031 | 0.519 | 0.035 | 0.043 | 0.593 |
| N-acetylaspartic acid | -0.014 | 0.030 | 0.718 | -0.008 | 0.031 | 0.887 | -0.013 | 0.042 | 0.873 |
| N-acetylhistidine | -0.033 | 0.029 | 0.384 | -0.051 | 0.030 | 0.161 | 0.074 | 0.041 | 0.178 |
| N-acetylmethionine | -0.014 | 0.030 | 0.724 | -0.005 | 0.031 | 0.913 | 0.039 | 0.043 | 0.540 |
| N-acetylornithine | 0.203 | 0.031 | <0.0001 | 0.274 | 0.031 | <0.0001 | 0.212 | 0.043 | 0.0002 |
| N-acetyltryptophan | 0.076 | 0.030 | 0.032 | 0.086 | 0.031 | 0.017 | 0.111 | 0.042 | 0.037 |
| N-alpha-acetylarginine | -0.011 | 0.031 | 0.782 | -0.066 | 0.032 | 0.088 | -0.017 | 0.044 | 0.822 |
| N-carbamoyl-beta-alanine | 0.096 | 0.030 | 0.005 | 0.102 | 0.030 | 0.003 | 0.104 | 0.041 | 0.049 |
| N-methylproline | 0.015 | 0.031 | 0.721 | 0.129 | 0.031 | 0.0002 | 0.086 | 0.043 | 0.136 |
| N1-acetylspermidine | 0.087 | 0.030 | 0.011 | 0.099 | 0.030 | 0.005 | 0.048 | 0.042 | 0.417 |
| N1-methyl-2-pyridone-5-carboxamide | 0.107 | 0.030 | 0.002 | 0.036 | 0.031 | 0.384 | 0.082 | 0.042 | 0.138 |
| N4-acetylcytidine | -0.096 | 0.031 | 0.007 | -0.061 | 0.031 | 0.107 | -0.153 | 0.043 | 0.004 |
| N6-acetyllysine | -0.034 | 0.031 | 0.387 | -0.055 | 0.031 | 0.155 | 0.020 | 0.043 | 0.794 |
| N6,N6-dimethyllysine | -0.045 | 0.032 | 0.263 | -0.024 | 0.033 | 0.596 | 0.008 | 0.045 | 0.917 |
| N6,N6,N6-trimethyllysine | -0.024 | 0.030 | 0.549 | -0.001 | 0.031 | 0.980 | 0.083 | 0.042 | 0.136 |
| niacinamide | 0.043 | 0.030 | 0.265 | -0.003 | 0.031 | 0.956 | 0.022 | 0.043 | 0.765 |
| NMMA | 0.189 | 0.031 | <0.0001 | 0.193 | 0.032 | <0.0001 | 0.127 | 0.044 | 0.020 |
| ornithine | 0.013 | 0.030 | 0.739 | 0.001 | 0.031 | 0.982 | -0.006 | 0.042 | 0.926 |
| pantothenate | 0.182 | 0.031 | <0.0001 | 0.140 | 0.032 | 0.0001 | 0.143 | 0.043 | 0.008 |
| phenylacetylglutamine | 0.014 | 0.030 | 0.722 | 0.009 | 0.031 | 0.879 | -0.082 | 0.042 | 0.138 |
| phenylalanine | 0.024 | 0.025 | 0.464 | 0.005 | 0.026 | 0.899 | -0.004 | 0.035 | 0.939 |
| phosphocholine | 0.088 | 0.030 | 0.013 | 0.037 | 0.031 | 0.376 | 0.106 | 0.043 | 0.050 |
| pipecolic acid | 0.218 | 0.031 | <0.0001 | 0.235 | 0.032 | <0.0001 | 0.177 | 0.044 | 0.002 |
| piperine | 0.098 | 0.031 | 0.007 | 0.098 | 0.032 | 0.008 | 0.086 | 0.044 | 0.138 |
| proline | -0.103 | 0.028 | 0.001 | -0.121 | 0.028 | 0.0001 | -0.060 | 0.039 | 0.262 |
| proline betaine | 0.0004 | 0.031 | 0.989 | 0.138 | 0.031 | 0.0001 | 0.096 | 0.043 | 0.084 |
| pseudouridine | -0.075 | 0.030 | 0.036 | -0.047 | 0.031 | 0.223 | -0.052 | 0.043 | 0.402 |
| ribothymidine | 0.026 | 0.031 | 0.539 | 0.024 | 0.032 | 0.585 | 0.027 | 0.044 | 0.708 |
| serine | 0.101 | 0.030 | 0.003 | 0.081 | 0.030 | 0.022 | 0.008 | 0.042 | 0.914 |
| solanidine | -0.081 | 0.031 | 0.027 | -0.067 | 0.032 | 0.082 | -0.040 | 0.044 | 0.540 |
| sphinganine | -0.032 | 0.030 | 0.402 | -0.031 | 0.031 | 0.459 | -0.015 | 0.043 | 0.848 |
| sphingosine | 0.019 | 0.031 | 0.641 | 0.063 | 0.032 | 0.102 | 0.056 | 0.043 | 0.369 |
| taurine | 0.078 | 0.030 | 0.027 | 0.088 | 0.031 | 0.014 | 0.032 | 0.042 | 0.631 |
| thiamine | 0.032 | 0.031 | 0.431 | 0.018 | 0.032 | 0.691 | -0.109 | 0.043 | 0.049 |
| threonine | -0.102 | 0.031 | 0.004 | -0.113 | 0.032 | 0.002 | -0.021 | 0.044 | 0.788 |
| thyroxine | -0.043 | 0.031 | 0.281 | -0.018 | 0.032 | 0.691 | -0.154 | 0.044 | 0.004 |
| trigonelline | 0.143 | 0.031 | <0.0001 | 0.134 | 0.032 | 0.0001 | 0.095 | 0.044 | 0.095 |
| trimethylamine-N-oxide | 0.112 | 0.031 | 0.001 | 0.126 | 0.031 | 0.0003 | 0.009 | 0.043 | 0.914 |
| trimethylbenzene | 0.074 | 0.031 | 0.044 | 0.093 | 0.032 | 0.012 | 0.132 | 0.043 | 0.014 |
| tryptophan | 0.024 | 0.030 | 0.549 | 0.011 | 0.030 | 0.830 | 0.004 | 0.042 | 0.942 |
| tyrosine | -0.022 | 0.028 | 0.579 | -0.035 | 0.029 | 0.361 | -0.069 | 0.040 | 0.202 |
| urate | 0.030 | 0.026 | 0.364 | 0.019 | 0.027 | 0.600 | 0.015 | 0.036 | 0.807 |
| urocanic acid | -0.024 | 0.031 | 0.561 | -0.038 | 0.032 | 0.362 | 0.013 | 0.044 | 0.878 |
| valine | 0.090 | 0.026 | 0.002 | 0.069 | 0.026 | 0.024 | 0.129 | 0.036 | 0.004 |
| vitamin A | 0.051 | 0.030 | 0.168 | 0.019 | 0.031 | 0.668 | 0.071 | 0.042 | 0.221 |
| xanthine | -0.065 | 0.030 | 0.072 | -0.050 | 0.031 | 0.185 | -0.091 | 0.042 | 0.099 |
| xanthopterin | -0.109 | 0.029 | 0.001 | -0.139 | 0.030 | <0.0001 | -0.036 | 0.041 | 0.577 |

Coefficients represent one SD change in dietary scores and peak relative VO_2_ per one SD increase in inverse rank normalized metabolite abundance. Model adjusted for sex, age, total caloric intake, BMI, smoking status, cholesterol, HDL, systolic blood pressure, hypertension medication use, diabetes, and physical activity index. P-values were adjusted for false discovery rate of 5% for each diet score and peak VO_2_ separately.
